# Environmental Chemical and Psychosocial Stress Exposures during Pregnancy and Child Behavioral Problems through 10 Years of Age in a South African Birth Cohort

**DOI:** 10.64898/2026.11.18.26368331

**Authors:** Sarina Abrishamcar, Stephanie M. Eick, Todd M. Everson, M. Daniele Fallin, Shakira F. Suglia, Jasmin Chovatiya, Ravikumar Jagani, Robert O. Wright, Syam S. Andra, Dana Boyd Barr, Nadia Hoffman, Heather J. Zar, Dan J. Stein, Anke Hüls

**Author notes:** Corresponding author Anke Hüls, PhD, MS. Deceased December 6^th^, 2025 (Dan J. Stein).

## Abstract

**Introduction:** Prenatal exposure to environmental chemicals and psychosocial stress often co-occur in low- and middle-income settings, yet their joint effects on child behavior from preschool to school-age remain unknown. We investigated the joint associations of prenatal chemical metabolites and psychosocial stress on child behavior in the first decade of life in the South African birth cohort, the Drakenstein Child Health Study.

**Methods:** Among 643 mother-child pairs, we quantified 25 chemical metabolites spanning nine classes (organophosphates, pyrethroids, neonicotinoid, chlorinated phenol, polycyclic aromatic hydrocarbons, parabens, bisphenols, UV filters, other biocides) in second-trimester maternal urine and characterized prenatal psychosocial stress as a five-indicator latent construct using confirmatory factor analysis. Internalizing and externalizing problems were assessed with the Child Behavior Checklist (CBCL) at 2, 3.5, 5, 6.5, 8, and 10 years. We fit adjusted generalized estimating equations for single exposures with false discovery rate correction. We assessed the joint effects of the positively associated exposure classes using quantile-based g-computation (QG-COMP). Models were fit separately in the preschool (2 to 5 years) and school-age (6.5 to 10 years) developmental windows. Finally, we tested effect measure modification by maternal psychosocial stress and by child sex.

**Results:** Chemical associations were confined to the preschool window. Three polycyclic aromatic hydrocarbon (PAH) metabolites were associated with increased externalizing problems (2-hydroxyfluorene 1.49 [95% CI 0.37, 2.60]; 1-naphthol 1.14 [0.21, 2.07]; 1-hydroxypyrene 1.05 [0.14, 1.95]), as was the PAH mixture (externalizing ψ=1.00 [0.32, 1.68]; internalizing ψ=0.95 [0.26, 1.64]). Psychosocial associations persisted across both windows (e.g. latent stress score and externalizing problems: 1.52 [0.83, 2.22] preschool, 1.45 [0.77, 2.14] school-age). The joint PAH and psychosocial mixture showed the largest effects at preschool (externalizing ψ=2.15 [1.15, 3.15]; internalizing ψ=2.02 [0.98, 3.06]), with psychosocial factors contributing most to the joint effect. Preschool associations were present among boys, but not girls (joint mixture, externalizing: ψ=3.42 [2.09, 4.69] in boys versus 0.74 [-0.67, 2.14] in girls, interaction p=0.004). There was no evidence of modification by maternal psychosocial stress.

**Discussion:** Prenatal urinary PAH metabolite levels and psychosocial exposures were associated with child behavioral problems through distinct developmental courses, with PAH associations limited to early childhood and psychosocial associations persisting into the school-age years. Their joint effect exceeded that of either domain alone, and was heightened among boys during the preschool years. These findings highlight the importance of joint chemical and non-chemical stressors on child mental health.

## Introduction

Pregnancy is a critical time window for neurodevelopment, as the developing fetus is uniquely susceptible to environmental toxicants and maternal psychosocial exposures. Prenatal exposure to environmental chemicals, such as pesticides, polycyclic aromatic hydrocarbons (PAHs), environmental phenols, or parabens, has been associated with child psychopathology, a broad range of emotional, behavioral, and cognitive disorders that can emerge from early life through adolescence^1–7^. Independently, prenatal psychosocial factors, such as maternal mental health and traumatic life events, can dysregulate the maternal-placental-fetal endocrine, immune, and metabolic systems, and have been linked to increased internalizing and externalizing behavioral problems in childhood ^8–18^. Responses within these systems are sexually dimorphic, and male and females may therefore differ in their susceptibility to both prenatal environmental and psychosocial exposures^19–21^. These exposures are highly prevalent in low-income communities and low- and middle-income countries (LMICs), where higher rates of poverty and structural inequalities disproportionately burden communities, leading to co-occurring chemical and psychosocial exposures that compound risk for adverse child neurodevelopmental outcomes ^22,23^.

While environmental toxicant exposure and psychosocial factors have traditionally been studied in isolation, growing evidence suggests that these exposures share overlapping biological pathways, including hypothalamic-pituitary-adrenal (HPA) axis dysregulation, oxidative stress, and neuroinflammation, and therefore may exhibit synergistic effects on child psychopathology ^24^. Additionally, psychosocial stressors may amplify the harmful effects of chemical toxicant exposure by lowering the developing brain’s threshold for neurotoxicity, such that children of mothers experiencing greater psychosocial burden may be disproportionately vulnerable to toxicant-associated harm ^17,24,25^. However, few studies have modeled the joint effects of prenatal chemical exposure and psychosocial factors on child behavioral outcomes ^26–30^. Assessing both joint effects and effect measure modification (EMM) offers complementary insights; joint effects capture the combined impact of co-occurring exposures on child psychopathology, whereas EMM identifies whether the magnitude of toxicant-associated harm differs by psychosocial stress level. Evaluating both allows for a more comprehensive understanding of how these exposures interact to shape child behavioral outcomes and can help identify both universal and targeted prevention strategies for vulnerable populations^31^.

Child behavior is typically assessed at a single time point or within a single developmental window, which cannot capture how exposure-associated risk emerges, persists, or attenuates across development^32^. Internalizing and externalizing problems change substantially across early and middle childhood, and the transition from preschool to school-age years brings change in social environments and in how behavior is expressed and reported^32–34^. Repeated assessments across childhood allow these patterns to be characterized and can help identify the developmental windows in which intervention may be most effective.

Here, we investigated the associations between prenatal urinary chemical metabolite levels and psychosocial stressors on child behavior assessed six times through10 years of age in the South African birth cohort, the Drakenstein Child Health Study (DCHS) and aimed to elucidate the role that maternal psychosocial stress plays, both as a co-exposure, and as an effect modifier of chemical exposure. We first assessed associations between 25 individual chemical metabolite levels, spanning nine chemical classes, and internalizing and externalizing problems across the preschool (2-5 years) and school-age (6.5-10 years) developmental windows, and tested whether these associations changed with time. We then used an environmental mixture method, quantile-based g-computation (QG-COMP), to estimate the joint effects of the chemical classes that were associated with child behavior in the single-exposure models. We modeled these classes alone and in combination with maternal psychosocial factors and identified the mixture components that contributed most to the joint effect. We characterized prenatal psychosocial stress as a multidimensional latent construct using confirmatory factor analysis to capture psychosocial burden across five indicators measured during the second trimester of pregnancy. Finally, we assessed whether these mixture associations differed by maternal psychosocial stress or by child sex.

## Methods

### Study population

The Drakenstein Child Health Study (DCHS) is a well-characterized, population-based birth cohort study based in a peri-urban region outside of Cape Town, South Africa. The cohort, which has previously been described in detail, is comprised of mother-child pairs from Black African and mixed races based in two peri-urban regions of Cape Town, South Africa. Consenting pregnant women (N=1137) were recruited during their second trimester of pregnancy from two primary health clinics, TC Newman and Mbekweni, between 2012 and 2015^35^. Mother-child pairs have been followed closely from birth up to at least 10 years. Ethical approval for human subjects research was given by the Human Research Ethics Committee of the Faculty of Health Sciences at the University of Cape Town (401/2009) and by the Western Cape Provincial Health Research committee. Written consent for participation was obtained from the mother on behalf of herself and the child, and mothers were reconsented annually in the mother’s preferred language: English, Afrikaans, or isiXhosa. Child assent was obtained annually from 7 years of age where the child was cognitively able to provide this.

### Child behavior

Child behavior was assessed at six time points : at 2, 3.5, and 5 years of age using the preschool version of the Child Behavior Checklist (CBCL/1.5-5) and at 6.5, 8, and 10 years of age using the school-age version of the Child Behavior Checklist (CBCL/6-18)^36,37^. Both versions are parent/caretaker report instruments designed to assess behavioral and emotional problems in children and are widely used in sub-Saharan Africa^38,39^. The CBCL/1.5-5 comprises 100 items and the CBCL/6-18 comprises 113 items. Items are scored based on a parent/caretaker’s assessment of how true each item is on a three-point Likert scale with 0 meaning “not true” and 2 meaning “very true or often true”. Both versions group items into age-appropriate syndrome scales (seven for the preschool version, eight for the school-age version), which in turn group into two higher-order subscales called internalizing problems and externalizing problems. Internalizing problems captures emotionally reactive, anxious/depressed, somatic, and withdrawn symptoms in CBCL/1.5-5 and anxious/depressed, withdrawn/depressed, and somatic complaints in CBCL/6-18. Externalizing problems capture attention problems and aggressive behavior in CBCL/1.5-5 and rule-breaking behavior and aggressive behavior in CBCL/6-18. For both subscales, a higher score indicates more behavioral symptoms.

To allow comparability across the two CBCL versions and across child ages, we used age- and sex-standardized T-scores (mean=50, SD=10) derived from the ASEBA normative sample as our outcomes. We modeled the internalizing and externalizing T-scores as continuous outcomes in all analyses.

### Prenatal urinary environmental chemical metabolites

Environmental chemical metabolites were measured in biobanked urine samples collected from mothers during the second trimester of pregnancy from 2012-2015. Urine samples were collected using a spot, clean catch method into sterile polypropylene collection tubes. Samples were transported, processed, aliquoted, and stored frozen in the DCHS research laboratory and curated in the DCHS biorepository using the Freezerworks inventory system. The biomarker measurements were provided by the Human Health Exposure Analysis Resource (HHEAR) Targeted Analysis Laboratory at the Icahn School of Medicine at Mount Sinai in New York, NY, using a previously developed and validated bioanalytical method^40^. A multiclass LC-MS/MS analytical method was used which included the quantification of eight organophosphate (OP) insecticide metabolites, one neonicotinoid insecticide metabolite, two pyrethroid insecticide metabolites, one chlorinated phenol, five polycyclic aromatic hydrocarbons (PAHs), three parabens, two bisphenols, two UV filters, and two biocides. Quality control consisted of experimental blanks (reagent-and matrix-based), matrix spikes at low, medium, and high validation levels, investigator-initiated paired duplicate samples blinded to the lab, two HHEAR QC urine pools (three of each analyzed per 100 samples), NIST standard reference materials 3672 and 3673, and archived, previously characterized proficiency-testing material, as outlined in HHEAR^41^.

### Prenatal psychosocial stress

Measures of psychosocial stress were obtained from mothers via self-report during the second trimester of pregnancy, and included depressive symptoms, psychological distress, traumatic life events, and physical and emotional interpersonal violence^42^. We used continuous scores for each psychosocial factor by summing the items within each scale to obtain a total score.

Depressive symptoms were measured using the Edinburgh Postnatal Depression (EPDS) scale, which is designed to measure postnatal depression^43^ and has also been verified for prenatal depression in this cohort ^44,45^. It consists of 10 items with scores ranging from 0 to 30, with higher scores indicating more severe depressive symptoms. Psychological distress was measured using the WHO-endorsed Self-Reporting Questionnaire (SRQ-20), which consists of 20 items with scores ranging from 0 to 20, with higher scores indicating higher levels of psychological distress ^44^.

Traumatic life events were measured using the World Mental Health Life Events Questionnaire, used to assess exposure to 13 areas of stressful or negative life events^46,47^. This instrument consists of 17 items with scores ranging from 0 to 17. Items were summed to obtain a total, continuous score.

Interpersonal violence (IPV) was assessed using the Intimate Partner Violence questionnaire ^48,49^ which captures lifetime and recent exposure to physical and emotional interpersonal violence. A continuous score was calculated by summing items in each domain, with a higher score indicating greater exposure to IPV.

### Confirmatory factor analysis of prenatal psychosocial stress

To characterize prenatal psychosocial stress as a multidimensional construct, we constructed a latent psychosocial stress variable using confirmatory factor analysis (CFA) implemented in the *lavaan* R package. A single latent stress factor was specified to underlie the five observed psychosocial factors as described above. The model was estimated using robust maximum likelihood (MLR) with full information maximum likelihood (FIML) to account for missing data. Based on conceptual overlap, residual correlations were specified between depressive symptoms (EPDS) and psychological distress (SRQ-20) and between physical and emotional IPV (IPV). Model fit was evaluated using standard indices, including the comparative fit index (CFI), Tucker-Lewis index (TLI), root mean square error of approximation (RMSEA), and standardized root mean square residual (SRMR).

Individual-level latent factor stress scores were estimated using the regression (empirical Bayes) method, which generates predicted latent values based on observed indicators and the estimated model parameters from the fitted CFA model. The continuous latent stress scores were dichotomized at the 75th percentile to define high versus low stress for downstream analyses. We also conducted sensitivity analyses using the latent stress score dichotomized at the median and as a continuous variable.

### Statistical Analysis

All analyses were restricted to participants with complete prenatal urinary environmental chemical metabolite measurements (n=643) Concentrations below the LOD were treated as left-censored and imputed using the mice.impute.leftcenslognorm function from the *qgcomp* R package, which draws from a left-censored log-normal distribution^50^.

Three metabolites with detection frequencies below 50% were dichotomized (detected vs. non-detected) in single-exposure models to stabilize estimates in the presence of a high proportion of non-detects. Chemical metabolite concentrations were natural log-transformed.

Non-normal psychosocial variables were natural log-transformed, and a constant of 1 was added to psychosocial survey scales that had zero-values.

All models were adjusted for self-reported race (Black African/Mixed), child sex, child age at CBCL assessment, maternal age at enrollment, and socioeconomic status (SES; sum composite score of maternal employment, maternal education, household income, and household assets), defined as the minimally sufficient set of confounders from a directed acyclic graph (DAG) (Figure S1). Models with PAH metabolites as exposure were additionally adjusted for maternal smoking as defined by second-trimester urine cotinine levels (non-smoker < 10 ng/ml, passive smoker (10-500 ng/ml), active smoker (> 500 ng/ml)) as many PAH metabolites are derived from tobacco smoke.^42^ Models with urinary chemical metabolites as exposure were additionally adjusted for prenatal urinary creatinine concentration to account for variation in urine dilution.

### Multiple imputation of missing values

Missing data were assumed to be missing at random (MAR), so we used multiple imputation by chained equations (MICE) using the *mice* R package to impute missing values in psychosocial factors, covariates, and CBCL outcome measures^51^. Thirty-five imputed datasets with 50 iterations were generated using predictive mean matching for continuous variables, left-censored log-normal for chemical metabolite concentrations below LOD, and polytomous regression for categorical variables. We iterated the main generalized estimating equation (GEE) models (single exposure models) across the 35 imputed datasets and pooled summary statistics using Rubin’s rules. Because of the complexity of downstream confirmatory factor analysis and mixture analyses that do not readily support pooling via Rubin’s rules and is computationally intensive, the average imputed dataset was used.

### Single exposure models

We conducted single-exposure adjusted GEE models to assess the associations between each prenatal environmental chemical metabolite or psychosocial factor and child internalizing and externalizing behavior across three time windows: the preschool waves (2, 3.5, 5 years), the school-age wave (6, 8, 10 years) and the full longitudinal model (all six waves) combined. GEE models were specified with an exchangeable working correlation structure, clustered on child ID to account for repeated CBCL measures. We adjusted for covariates as described above and we additionally adjusted the full longitudinal model for CBCL version (preschool vs school-age) to account for differences between the two instruments. All GEE estimates were pooled across imputations using Rubin’s rules. We adjusted for multiple testing using the Benjamini-Hochberg False Discovery Rate (BH-FDR) procedure, applied separately for each outcome and developmental window.

### Time-varying single-exposure models

To test whether the association between prenatal exposures and child behavior changed over the course of childhood, we re-fit each single-exposure GEE model with interactions between the exposure and both linear and quadratic terms for child age i.e. *exposure x (age+age*^2^). Weincluded a quadratic term to allow for non-linear effects over time. Time-varying models were conducted for exposure classes in which one or more individual exposures were positively associated with either outcome in the primary single-exposure models. Models were fit across all six assessments and separately within the preschool and school-age windows. Within each imputation dataset, we tested the joint significance of the linear and quadratic interactions terms via a 2-degree-of-freedom Wald test and computed the marginal exposure effect at each assessment age. We pooled the joint Wald statistic across the 35 imputations using the D2 procedure^52^, and pooled the age-specific marginal effect estimates and standard errors using Rubin’s rules. We applied the BH-FDR correction across the D2-pooled interaction p-values separately for each outcome and developmental window.

### Mixture models

We utilized quantile-based g-computation (QG-COMP), an environmental mixture method to assess the joint effects of prenatal chemical or psychosocial mixtures on CBCL scores at the preschool and school-age developmental windows^50^. QG-COMP estimates the effect of simultaneously increasing all exposures in the mixture by one quintile on CBCL scores and allows for exposures to have varying directions of effect. QG-COMP models were fit using 500 bootstrap samples, and 95% confidence intervals were obtained from the bootstrap percentile distribution. QG-COMP exposure weights were obtained from non-bootstrapped models. Exposure classes in which one or more individual exposures were positively associated with either outcome in the single exposure models were included in the QG-COMP modeling.

In secondary analyses, we expanded QG-COMP models to include psychosocial factors as co-exposures to assess the joint effects of prenatal chemical metabolites and psychosocial factors on CBCL outcomes. All models were adjusted as described above.

### Effect measure modification

To assess whether the association between a chemical class mixture and CBCL scores differed by maternal psychosocial stress, we conducted effect measure modification (EMM) analyses using the dichotomized latent stress score (high stress vs low stress; at 75^th^ percentile) as a modifier. An interaction term was included between the chemical mixture index and the latent stress score in the QG-COMP models. We also conducted a sensitivity analysis using a latent stress score dichotomized at the median and as a continuous variable.

Previous studies have suggested that sensitivity to prenatal environmental insults may differ by child sex, reflecting sexually dimorphic placental function and fetal programming^19–21^. To assess whether the association between an exposure class mixture and CBCL scores differed by child sex, we conducted EMM analyses. An interaction term was included between the chemical mixture index and child sex in the QG-COMP models.

### Quantitative bias analysis for exposure measurement error

We conducted quantitative bias analysis (QBA) for exposure measurement error in the urinary PAH metabolites, the exposure class for which the single-exposure models identified positive associations with behavior problems. Urinary PAH metabolites have short half-lives, so a single spot sample likely characterizes recent exposure rather than average gestational exposure^53^. Under a classical, non-differential error assumption, the observed regression coefficient is the true coefficient multiplied by the reliability of the exposure measure. Therefore, the bias-adjusted coefficient is the observed coefficient divided by the reliability, in this case the intraclass correlation coefficient (ICC) of repeated measurements within an individual^54–56^ (Equation 1).

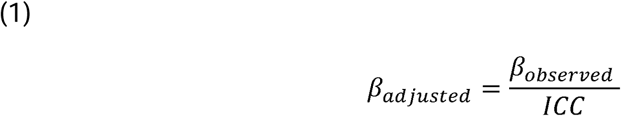

Since repeated urine samples were not collected during pregnancy, the ICC cannot be estimated internally and were instead transported from external studies. We identified three previous studies that reported ICCs for repeated urinary PAH metabolites across pregnancy. Puerto Rico Testsite for Exploring Contamination (PROTECT), a Puerto Rico birth cohort, collected three spot urines between 16 and 28 weeks of gestation from 659 women and reported ICCs of 0.39 to 0.56 across the five metabolites, standardized for specific gravity^57^. NYU Children’s Health and Environment Study (NYU CHES), a New York City cohort, collected three spot urines at approximately 10, 20, and 30 weeks gestation from 671 women and reported ICCs of 0.41 to 0.58 across the five metabolites, standardized for creatinine^58^. Third, a three-site study within the ECHO Cohort reported the site-specific of ICCs of 0.07 to 0.79 from 30 participants per site, using either spot urine or first morning voids^59^. PROTECT and NYU CHES agreed to within 0.04 for every metabolite, despite differing in population and in method of dilution standardization. (Table S11).

We then treated the ICC as a bias parameter and conducted a multi-dimensional bias analysis,^60^ sweeping across the range of ICCs previously reported for urinary PAH metabolites during pregnancy (0.20 to 0.70). At each value, the regression coefficient and it’s confidence limits were divided by the ICC^56,61^. The ICC was treated as fixed at each value, so the confidence limits reflect sampling variability in the observed effect estimates alone and do not incorporate uncertainty in the externally derived ICCs. Therefore, the correction applies to the magnitude of effect rather than on the precision of effect estimates. We applied the correction to single-metabolite estimates only.

## Results

### Study population characteristics

The analytic sample included 643 mother-child pairs with complete prenatal urinary chemical metabolite measurements (Table 1). Approximately 47% of children were female, and the cohort was evenly split between Black African (50%) and Mixed race (50%). The mean maternal age at enrollment was 26.8 years (SD: 5.95). In terms of socioeconomic indicators, over half of mothers had not completed secondary education, over 85% of households earned less than R5,000 (South African rand; equivalent to 300 USD) per month, and approximately half of mothers were unemployed. For context, the lower- and upper-bound poverty line (per person per month) in 2025 was R1,415 to R2,846 (equivalent to 85 USD to 170 USD)^62^. Prenatal tobacco exposure was common, with approximately 43% of women classified as passive smokers and 36% as active smokers, while 14% reported any prenatal alcohol use. Prenatal psychosocial stress was highly prevalent in this cohort, with 22% of mothers above the threshold for psychological distress and 26% of mothers above the threshold for depression symptoms. Additionally, 72% of mothers reported experiencing at least 1 traumatic life event and half of mothers reported experiencing physical or emotional IPV during their lifetime. Missing data were low across all psychosocial measures (<7% for all measures).

**Table 1.** Population characteristics and psychosocial factors. Drakenstein Child Health Study, Cape Town, South Africa, 2012-2015.

| **A. Study Characteristics** |  |
|---|---|
| **N** | 643 |
| **Child Sex** |  |
| Female | 303 (47.1%) |
| Male | 340 (52.9%) |
| **Race** |  |
| Black African | 319 (49.6%) |
| Mixed Race | 324 (50.4%) |
| **Maternal age (years)** |  |
| Mean (SD) | 26.8 (5.95) |
| **Gestational age (weeks)** |  |
| Mean (SD) | 38.6 (2.31) |
| **Education** |  |
| Primary | 57 (8.9%) |
| Some secondary | 343 (50.2%) |
| Completed secondary | 208 (32.3%) |
| Any tertiary | 35 (5.4%) |
| **Household income** |  |
| <R1000/month | 238 (37.0%) |
| R1000-5000/month | 323 (50.2%) |
| >R5000/month | 82 (12.8%) |
| **Parental employment** |  |
| Unemployed | 331 (51.5%) |
| Employed | 312 (48.5%) |
| **SES Sum Score** |  |
| Median (IQR) | 0.2 (-1.2, 1.7) |
| **Prenatal tobacco exposure (maternal cotinine)** |  |
| Non-smoker (<10 ng/mL) | 133 (20.7%) |
| Passive smoker (10-499 ng/mL) | 273 (42.5%) |
| Active smoker (≥ 500 ng/mL) | 231 (35.9%) |
| **Prenatal alcohol exposure (ASSIST)** |  |
| Median (IQR) | 0 (0, 0) |
| Above threshold (ASSIST ≥ 4) | 197 (32%) |
| Missing | 27 (4.2%) |
| **B. Psychosocial Factors** |  |
| **Depressive symptoms (EPDS)** |  |
| Median (IQR) | 9.0 (6.0,13.0) |
| Above threshold (EPDS ≥13) | 160 (26%) |
| Missing | 24 (3.7%) |
| **Psychological distress (SRQ-20)** |  |
| Median (IQR) | 4.0 (1.0, 7.0) |
| Above threshold (SRQ-20 ≥ 8) | 136 (22%) |
| Missing | 26 (4.0%) |
| **Emotional interpersonal violence** |  |
| Median (IQR) | 5.0 (4.0, 7.0) |
| Any lifetime exposure | 313 (51%) |
| Missing | 25 (3.9%) |
| **Physical interpersonal violence** |  |
| Median (IQR) | 5.0 (5.0, 7.0) |
| Any lifetime exposure | 306 (50%) |
| Missing | 25 (3.9%) |
| **Traumatic life events (LEQ)** |  |
| Median (IQR) | 1.0 (0, 3.0) |
| At least 1 life event | 431 (72%) |
| Missing | 41 (6.4%) |
| **Latent Psychosocial Stress Score** |  |
| Median (IQR) | -0.6 (-1.8, 1.0) |
| High Stress (≥ 75th percentile) | 161 (25.0%) |

Most chemical metabolites were detected in more than 50% of participants. The metabolites 6-chloronicotinic acid (CINA-6), bisphenol S (BPS), and 2,4-dichlorophenol (2,4-DCP) had the lowest detection rates and were therefore dichotomized as detected vs. non-detected in statistical analysis (Table 2). Chemical metabolites were all positively correlated with one another, most strongly within classes, with the strongest correlations observed among the PAH metabolites (r=0.43 to 0.71), between the two UV filters benzophenone-1 (BP1) and oxybenzone (BP3) (r=0.80), and between the two pyrethroid metabolites trans-3-(2,2-dichlorovinyl)-2,2-dimethylcyclopropane-l-carboxylic acid (TDCCA) and 3-phenoxybenzoic acid (PBA) (r=0.66) (Figure S2).

**Table 2:** Distribution of prenatal urinary environmental chemical metabolites measured at the second trimester of pregnancy, Drakenstein Child Health Study, Cape Town, South Africa, 2012-2015. Shown are the percentage of samples above the limit of detection (% ≥ LOD), geometric mean (GM) and geometric standard deviation (GSD), and the chemical concentrations at the 25th percentile, median, and 75th percentile. Values below to LOD are indicated as <LOD. Concentrations are reported in ng/mL unless otherwise specified.

| Metabolite | % ≥<br>LOD | GM (GSD) | Median [IQR] |  |  |
|---|---|---|---|---|---|
|  |  |  | 25th | Median | 75th |
| **Organophosphates (OPs)** |  |  |  |  |  |
| diethyl phosphate (DEP) | 94% | 2.61 (3.00) | 1.38 | 2.67 | 4.78 |
| diethyl thiophosphate (DETP) | 82% | 0.96 (2.95) | 0.44 | 0.9 | 1.64 |
| dimethyl dithiophosphate (DMDP) | 54% | 0.45 (3.24) | <LOD | 0.46 | 0.93 |
| dimethyl phosphate (DMP) | 98% | 10.85 (3.42) | 4.74 | 11.7 | 23.45 |
| dimethyl thiophosphate (DMTP) | 98% | 6.08 (2.29) | 3.31 | 5.49 | 9.99 |
| p-nitrophenol (PNP) | 73% | 0.95 (2.65) | <LOD | 0.89 | 1.65 |
| 3,5,6-trichloro-2-pyridinol (TCPy) | 95% | 2.57 (2.74) | 1.31 | 2.66 | 5.04 |
| **Pyrethroids** |  |  |  |  |  |
| 3-phenoxybenzoic acid (PBA) | 60% | 0.53 (2.92) | <LOD | 0.55 | 1.08 |
| trans-3-(2,2-dichlorovinyl)-2,2-dimethylcyclopropane-l-carboxylic acid (TDCCA) | 53% | 0.44 (3.06) | <LOD | 0.34 | 0.83 |
| **Neonicotinoid** |  |  |  |  |  |
| 6-chloronicotinic acid (CINA-6) | 35% | 0.22 (3.23) | <LOD | <LOD | 0.43 |
| **Chlorinated phenol** |  |  |  |  |  |
| 2,4-dichlorophenol (2,4-DCP) | 45% | 0.19 (3.02) | <LOD | <LOD | 0.37 |
| **Polycyclic aromatic hydrocarbons (PAHs) (ng/L)** |  |  |  |  |  |
| 2-hydroxyfluorene (2-FLUO) | 89% | 574.13 (2.81) | 300 | 590 | 1155 |
| 1-hydroxynaphthalene (1-NAP) | 90% | 1212.11 (3.31) | 510 | 1020 | 2580 |
| 2-hydroxynaphthalene (2-NAP) | 100% | 7584.3 (3.08) | 3340 | 7750 | 17200 |
| 1-hydroxyphenanthrene (1-PHEN) | 81% | 430.94 (3.01) | 200 | 450 | 850 |
| 1-hydroxypyrene (1-PYR) | 84% | 398.72 (2.39) | 220 | 380 | 645 |
| **Parabens** |  |  |  |  |  |
| Methyl paraben (MEPB) | 100% | 92.18 (5.75) | 30.45 | 111 | 345.5 |
| Ethyl paraben (ETPB) | 71% | 1.56 (5.32) | <LOD | 0.95 | 3.8 |
| Propyl paraben (PRPB) | 99% | 28.6 (6.4) | 8.63 | 34 | 117 |
| **Bisphenols** |  |  |  |  |  |
| Bisphenol A (BPA) | 96% | 2.05 (2.39) | 1.13 | 1.95 | 3.56 |
| Bisphenol S (BPS) | 37% | 0.36 (4.03) | <LOD | <LOD | 0.89 |
| **UV filters** |  |  |  |  |  |
| Benzophenone-1 (BP1) | 55% | 0.7 (4.46) | <LOD | 0.58 | 1.38 |
| Oxybenzone (BP3) | 97% | 6.55 (3.66) | 3.04 | 5.17 | 10.6 |
| **Other biocides** |  |  |  |  |  |
| Pentachlorophenol (PCP) | 69% | 0.39 (3.12) | <LOD | 0.38 | 0.8 |
| Triclosan (TCS) | 93% | 8.56 (4.84) | 3.09 | 6.67 | 22.65 |

LOD=limitation of detection; GM=geometric mean; GSD=geometric standard deviation

Children completed a median of five of six possible CBCL assessments, and 99% contributed at least three. Wave-specific completion ranged from 417 children (65%) at 24 months to 630 (98%) at 10 years (Table S1). Mean internalizing and externalizing t-scores fell below the normative mean of 50 at every wave, and boys scored higher relative to US norms than girls at most waves (Figure S3).

### Confirmatory factor analysis for prenatal psychosocial stress

The CFA model demonstrated good fit (comparative fit index (CFI)=0.995, Tucker-Lewis index (TLI)=0.982, root mean square error of approximation (RMSEA)=0.052, standardized root mean square residual (SRMR)=0.012). All five indicators loaded positively onto the latent psychosocial stress factor, with standardized factor loadings ranging from 0.43 to 0.69 (Figure S4). The strongest contributions to the latent stress construct were observed for psychological distress (SRQ; loading = 0.69) and traumatic life events (LEQ; loading = 0.65), followed by emotional IPV (IPV; loading = 0.53). Moderate loadings were observed for prenatal depressive symptoms (EPDS; 0.46) and physical IPV (IPV; 0.43). The latent stress score was positively correlated with all five observed psychosocial indicators, with the highest correlations observed for psychological distress (r=0.84), traumatic life events (r=0.79), and emotional IPV (r=0.65), and the lowest for depressive symptoms (r=0.56) and physical IPV (r=0.52), consistent with the factor loading pattern (Figure S5). The distribution of the latent stress score was right-skewed, with the 75th percentile score being 1.00 (range: −3.42 to 11.62) (Figure S6).

### Single-exposure associations between prenatal exposures and longitudinal CBCL scores

We conducted single-exposure GEE models between each prenatal environmental chemical urinary metabolite or psychosocial factor and CBCL internalizing and externalizing T-scores across three time windows: all six waves (2-10 years), preschool waves (2, 3.5, 5 years), and school-age waves (6.5, 8, 10 years) (Table S2).

Across all six waves, only associations between prenatal psychosocial factors and CBCL scores reached significance after adjusting for covariates as described above and multiple testing correction (q < 0.05) (Figure S7). The latent stress score was associated with increased externalizing (β=1.49; 95% CI: [0.97, 2.00]) and internalizing (1.30 [0.81, 1.80]) problems per one IQR increase in the natural log-transformed exposure. Among individual psychosocial factors, psychological distress showed the strongest association with externalizing problems (1.87 [1.08, 2.65]), followed by traumatic life events (1.58 [0.60, 2.55]), emotional IPV (1.19 [0.54, 1.84]), and depressive symptoms (0.84 [0.35, 1.32]). Physical IPV was positively associated with externalizing problems but did not survive multiple testing correction (0.56 [0.11, 1.00], q=0.072).

For internalizing problems, all five individual maternal psychosocial stressors survived multiple testing correction. Maternal traumatic life events showed the strongest association (1.67 [0.79, 2.54]), followed by psychological distress (1.49 [0.76, 2.22]), emotional IPV (0.96 [0.36, 1.56]), depressive symptoms (0.73 [0.31, 1.14]), and physical IPV (0.66 [0.24, 1.08]). No environmental chemical metabolite was associated with longitudinal CBCL scores across all six waves at q < 0.05. The PAH metabolite 1-hydroxynaphthalene (1-NAP) was associated with increased externalizing problems at a suggestive threshold of q < 0.10 (0.77 [0.11, 1.42], q=0.095).

Stratifying by developmental window revealed that chemical associations were confined to the preschool period (Figure 1). One PAH metabolite, 2-hydorxyfluorene (2-FLUO) was associated with increased externalizing problems during the preschool waves after adjusting for covariates as described above and multiple testing correction (q < 0.05) (1.49 [0.37, 2.60]). Two further PAH metabolites were associated with increased externalizing problems at a suggestive threshold of q < 0.10: 1-NAP (1.14 [0.21, 2.07], q=0.070), and 1-hydroxypyrene (1-PYR) (1.05 [0.14, 1.95], q=0.092). Associations with internalizing problems were consistent in direction but did not survive multiple testing correction (2-FLUO: 1.15 [-0.06, 2.36], q=0.21; 1-PYR: 0.96 [0.07, 1.84], q=0.19). The latent stress score, in addition to four of the five psychosocial factors was associated with increased internalizing and externalizing problems at preschool. The association was strongest for psychological distress (externalizing: 2.21 [1.24, 3.19]) and the latent stress score (externalizing 1.52 [0.83, 2.22]; internalizing: 1.32 [0.56, 2.08]).

**Figure 1.**
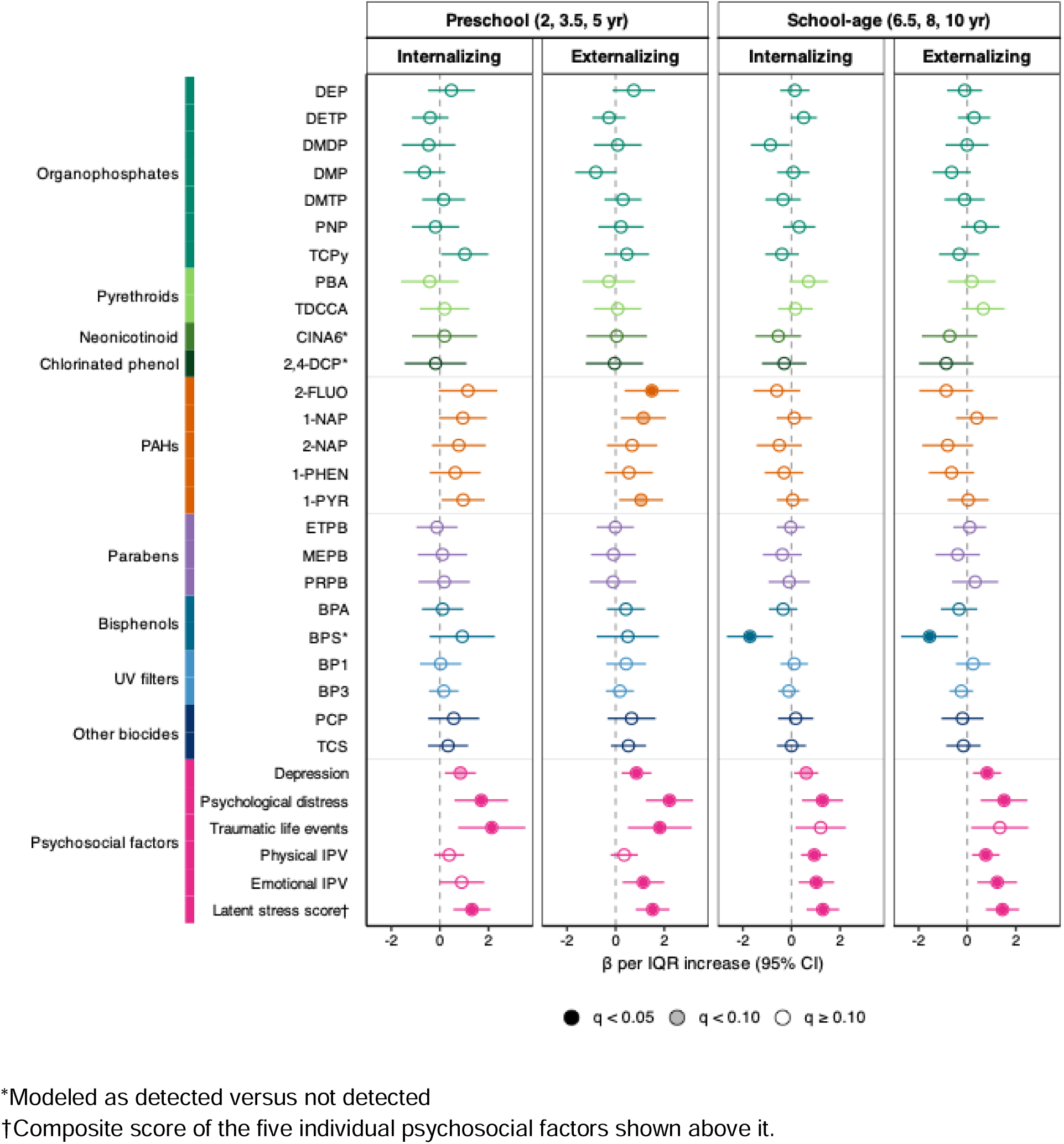
Single-exposure associations between prenatal environmental or psychosocial exposures and child internalizing and externalizing problems at preschool and school-age windows. Estimates are from GEE models with an exchangeable working correlation, fit across 35 multiply imputed datasets and pooled by Rubin’s rules. Preschool comprises the 2, 3.5, and 5 year assessments and school-age comprises the 6.5, 8, and 10 year assessments. Point estimates denote the change in CBCL T-score per one interquartile range (IQR) increase in the exposure, with horizontal lines denoting the corresponding 95% confidence intervals. The colored bar groups the exposure into ten exposure classes. The BH-FDR correction was applied across all exposures within each outcome and developmental window. Point estimate fill indicates the FDR-corrected q-value: filled q <0.05, gray q >0.10, and open q ≥ 0.10. All models were adjusted for self-reported race, child sex, child age at CBCL assessment, maternal age at enrollment, and SES. Models for urinary chemical metabolites were additionally adjusted for maternal urinary creatinine concentration and models for PAH metabolites were further adjusted for maternal smoking.

In the school-age waves, psychosocial associations persisted at comparable magnitude to the preschool period, with the latent stress score associated with a 1.45 point (0.77, 2.14) increase in externalizing and a 1.29 point (0.62, 1.97) increase in internalizing problems (Figure 1). Physical IPV, which was not associated with either outcome during the preschool waves, was associated with both internalizing (0.94 [0.40, 1.48]) and externalizing (0.77 [0.19, 1.34]) problems at school-age. In contrast, PAH associations were null in this window. Bisphenol S (BPS) detection was inversely associated with both internalizing (−1.71 [−2.66, −0.76]) and externalizing (−1.55 [-2.72, −0.38]) problems at school-age, while it was positively associated but not statistically significant at preschool.

### Time-varying single exposure associations between prenatal exposures and CBCL scores

To assess whether exposure effects changed across childhood, we re-fit the single-exposure GEE models with interactions between the exposure and both linear and quadratic terms for child age, across all six waves and within the preschool and school-age developmental windows. We conducted time-varying models for the PAH and psychosocial exposure classes, as they exhibited at least one individual exposure positively associated with either outcome in the primary single-exposure models. After adjusting for covariates, no exposure showed evidence of a time-varying effect surviving correction for multiple testing in any window (Figure S8; Table S3-S4). During the school-age waves, the latent stress showed suggestive evidence of change across waves for internalizing problems, but did not survive multiple testing correction (q=0.097). The association was null at 78 months (latent stress: 0.34 [−0.50, 1.17] per IQR) and emerged by 96 months (1.88 [0.88, 2.87]), remaining elevated at 120 months (1.66 [0.64, 2.68]).

### Prenatal PAH mixture

We assessed the joint effect of the five prenatal PAH metabolites on CBCL scores using QG-COMP (Figure 2; Table S5) at the preschool and school-age developmental windows. Consistent with the single-exposure models, the PAH mixture was associated with behavioral problems during the preschool period. A one-quintile increase in the PAH mixture was associated with a 1.00 point (95% CI: 0.32, 1.68) increase in externalizing problems and a 0.95 point (95% CI: 0.26, 1.64) increase in internalizing problems across the preschool waves. Because the overall preschool mixture effects were positive, we report the positive component weights, which represents each exposure’s contribution to the joint effect. In the preschool models, 1-NAP was the largest contributor to the joint effect for both internalizing and externalizing problems during preschool, followed by 2-FLUO and 1-PYR (Table S6).

**Figure 2:**
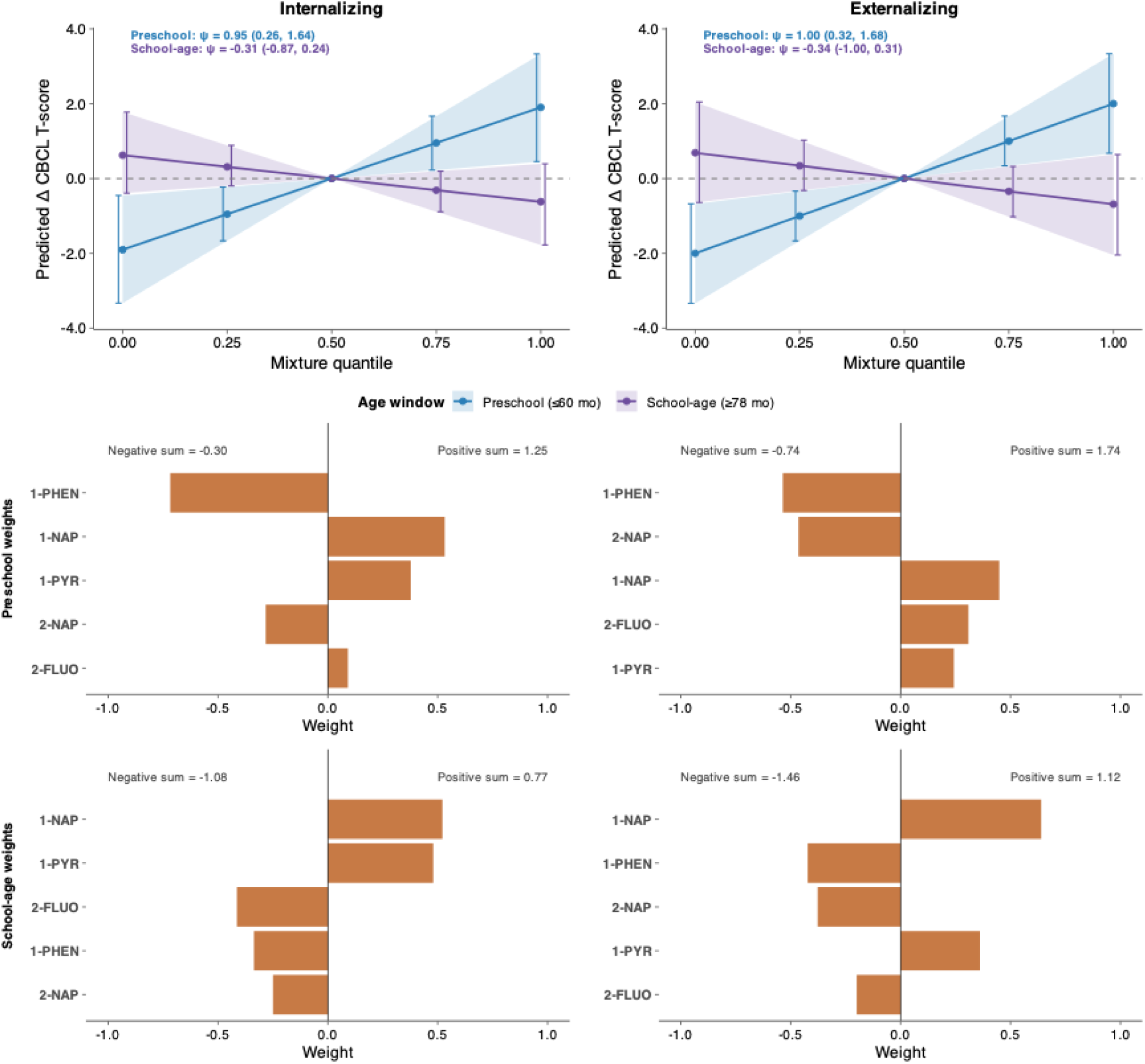
Quantile g-computation (QG-COMP) mixture associations between the prenatal PAH mixture and child internalizing and externalizing behavior at the preschool and school-age windows. Columns show internalizing (left) and externalizing (right) problems. Top row: QG-COMP marginal dose response curves for preschool (blue) and school-age (purple) developmental windows, showing the predicted change in CBCL T-score per one quintile increase in all exposures simultaneously, relative to the middle quintile, along with 95% confidence intervals. The estimated cumulative mixture effect (ψ) and 95% confidence intervals are denoted within each panel. Middle and bottom rows: QG-COMP mixture weights for the preschool and school-age windows, indicating the relative contribution and direction of each exposure to the overall mixture effect. Weights are normalized and therefore sum to 1 within each direction. The positive sum and negative sum are the summed effects of the positively and negatively weighted components, with their net equivalent to ψ. All models were adjusted for self-reported race, child sex, child age at CBCL assessment, maternal age at enrollment, maternal smoking, maternal urinary creatinine concentration, and SES.

The mixture was not associated with either outcome during the school-age waves (externalizing: ψ=−0.34, 95% CI: −1.00, 0.31; internalizing: ψ=−0.31, 95% CI: −0.87, 0.24).

### Prenatal psychosocial stress mixture

In contrast to the PAH mixture, the psychosocial stress mixture was associated with both outcomes in both developmental windows (Figure S9; Table S5). Across the preschool waves, a one quintile increase in the psychosocial mixture was associated with a 1.35 point (95% CI: 0.54, 2.16) increase in internalizing problems and a 1.43 point (95% CI: 0.62, 2.23) increase in externalizing problems. Estimates were comparable during the school-age waves: (internalizing: 1.59 [0.80, 2.39]; externalizing: 1.53 [0.64, 2.42]). For internalizing problems, the largest contributors to the joint effect were maternal traumatic life events or emotional IPV at preschool, and maternal physical and emotional IPV at school-age. For externalizing problems, the largest contributors to the joint effect were maternal emotional IPV and psychological distress at preschool, and maternal emotional IPV and depression at school-age (Table S6).

### Joint mixture of PAHs and psychosocial stress

When PAH metabolites and psychosocial factors were modeled together, the joint mixture was associated with both outcomes during the preschool wave and with internalizing problems at school-age (Figure 3; Table S5). At preschool, the magnitudes of effect were larger than either the PAH-only or psychosocial factor-only mixtures alone. The joint associations were strongest during the preschool waves, where a one quintile increase in the mixture was associated with a 2.15 point (95% CI: 1.15, 3.15) increase in externalizing problems and a 2.02 point (95% CI: 0.98, 3.06) increase in internalizing problems. Associations attenuated during the school-age wave, where the mixture remained associated with internalizing problems (ψ=1.16, 95% CI: 0.16, 2.16) but was no longer associated with externalizing problems (ψ=1.06, 95% CI: −0.05, 2.16). Traumatic life events, emotional IPV and physical IPV were the largest psychosocial contributors at preschool internalizing, preschool externalizing, and school-age internalizing respectively, and 1-NAP was the largest chemical contributor across both developmental windows and outcomes (Table S6).

**Figure 3:**
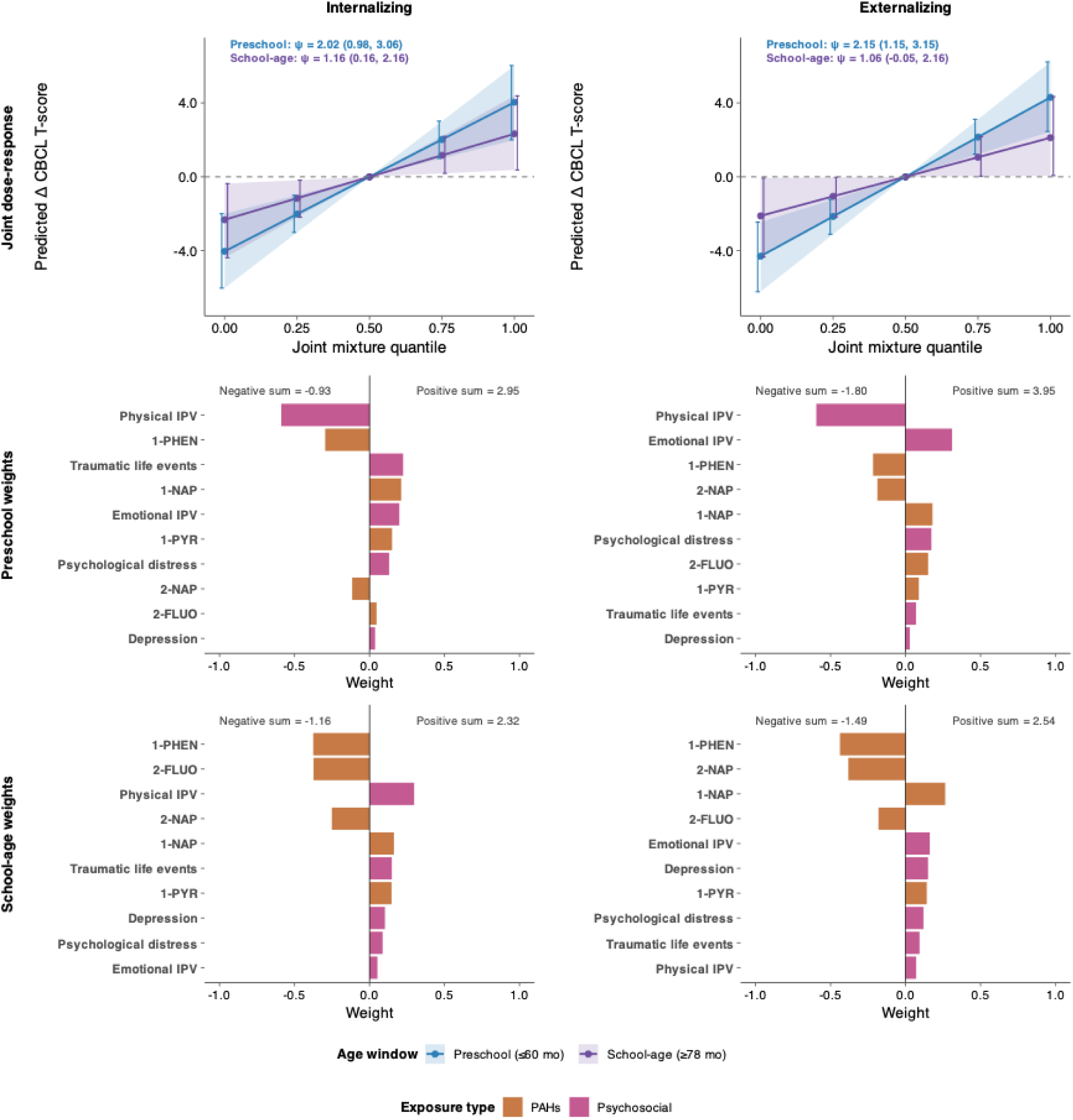
Quantile g-computation (QG-COMP) mixture associations between the joint prenatal PAH and psychosocial stress mixture and child internalizing and externalizing behavior at the preschool and school-age windows. Columns show internalizing (left) and externalizing (right) problems. Top row: QG-COMP marginal dose response curves for preschool (blue) and school-age (purple) developmental windows, showing the predicted change in CBCL T-score per one quintile increase in all exposures simultaneously, relative to the middle quintile, along with 95% confidence intervals. The estimated cumulative mixture effect (ψ) and 95% confidence intervals are denoted within each panel. Middle and bottom rows: QG-COMP mixture weights for the preschool and school-age windows, indicating the relative contribution and direction of each exposure to the overall mixture effect. Weights are normalized and therefore sum to 1 within each direction. The positive sum and negative sum are the summed effects of the positively and negatively weighted components, with their net equivalent to ψ. PAH exposure weights are shown in orange and psychosocial exposure weights are shown in pink. All models were adjusted for self-reported race, child sex, child age at CBCL assessment, maternal age at enrollment, maternal smoking, maternal urinary creatinine concentration, and SES.

### Effect modification by maternal psychosocial stress

We assessed whether the association between the prenatal PAH mixture and CBCL scores differed by prenatal maternal psychosocial stress, dichotomized at the 75^th^ percentile of the latent stress score (Figure 4). During the preschool waves, stratum-specific estimates were positive among children of mothers who experienced low stress low-stress mothers (externalizing: ψ=1.19, 95% CI: 0.47, 1.92, p=0.001; internalizing: ψ=1.09, 95% CI: 0.35, 1.83, p=0.004) and attenuated among children of mothers who experienced high stress (externalizing: ψ=0.51, 95% CI: −0.54, 1.54, p=0.38; internalizing: ψ=0.55, 95% CI: −0.54, 1.62, p=0.37), but neither interaction was statistically significant (p=0.24 and p=0.35, respectively) and the stratum-specific confidence intervals overlapped substantially. During the school-age waves, the mixture was not associated with either outcome among children of mothers who experienced low stress (externalizing: ψ=−0.02, 95% CI: −0.73, 0.69, p=0.96; internalizing: ψ=−0.26, 95% CI: −0.84, 0.32, p=0.37), but was inversely associated with externalizing problems among children of mothers who experienced high stress (ψ=−1.17, 95% CI: −2.18, −0.18, p=0.028; interaction p=0.044). The corresponding internalizing estimate was not statistically significant. This was the only interaction to reach nominal significance across the four models tested.

**Figure 4.**
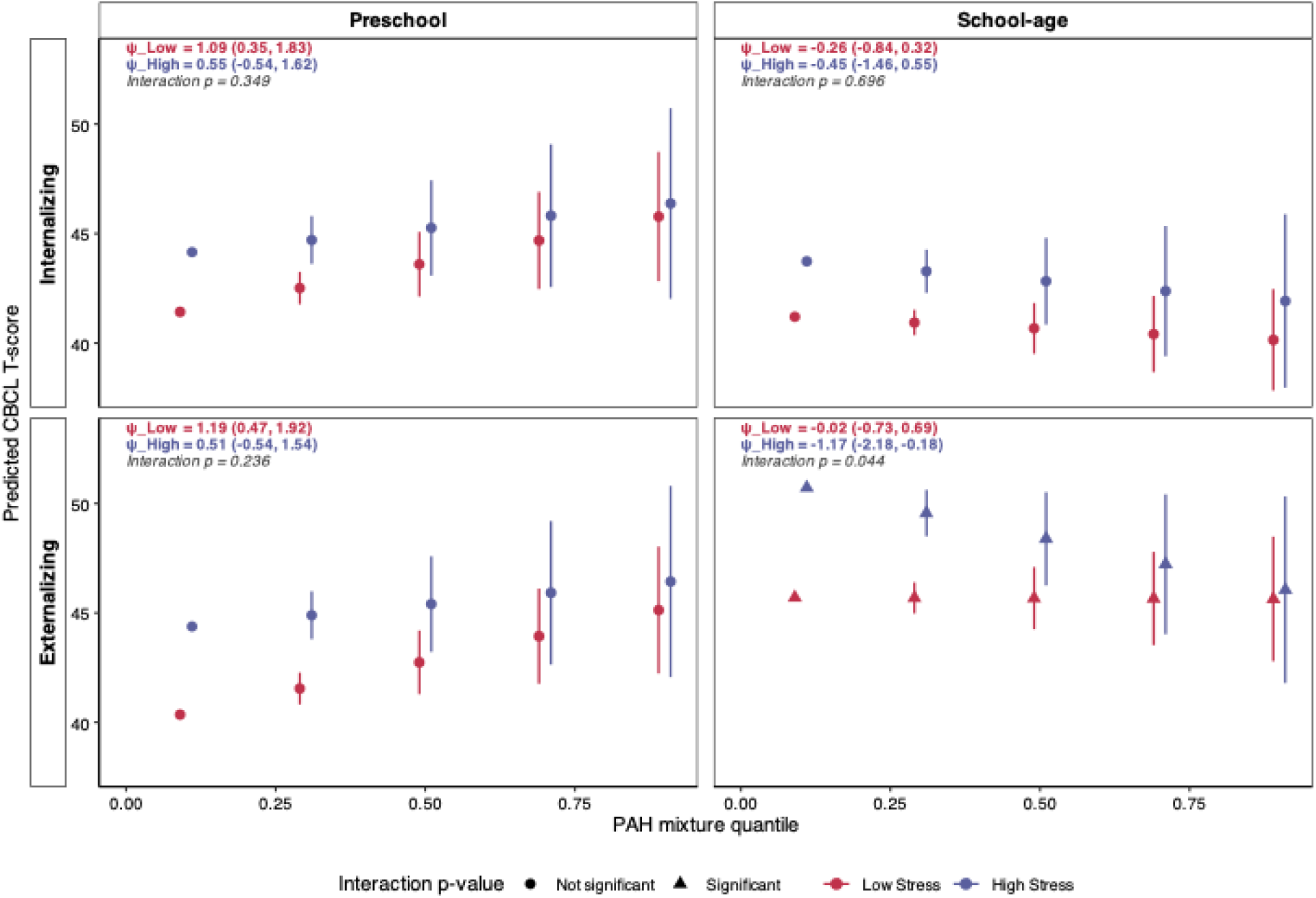
Effect measure modification by maternal latent psychosocial stress score of the association between the prenatal PAH mixture and child internalizing (top row) and externalizing (bottom row) problems in the preschool (left column) and school-age (right column) windows. Stress was dichotomized at the 75^th^ percentile of the latent stress score. Stratum-specific QG-COMP estimates, stratified by low (pink) and high (purple) maternal stress. Points show the predicted CBCL T-score at each quintile of the PAH mixture and vertical lines are the corresponding 95% confidence interval. Point shape indicates whether the mixture by stress interaction p-value was statistically significant, with a triangle shape indicating statistical significance (p < 0.05). The estimated cumulative mixture effect (ψ) for each stratum, 95% confidence intervals, and interaction p-value are denoted within each panel. Ψ is the change in CBCL T-score per one quintile increase in all exposures simultaneously. All models were adjusted for self-reported race, child sex, child age at CBCL assessment, maternal age at enrollment, maternal smoking, maternal urinary creatinine concentration, and SES.

This interaction was not robust to how psychosocial stress was parameterized. When the latent stress score was dichotomized at the median, the inverse school-age externalizing association was no longer present among children of mothers who experienced high stress (ψ=−0.27, 95% CI: −1.10, 0.62, p=0.54; interaction p=0.83), and when the score was modeled continuously the interaction was similarly attenuated (−0.11 per one-unit increase in the stress score, 95% CI: −0.29, 0.06; interaction p=0.19). In the preschool waves, the interaction using either the median split or continuous score was robust to the primary analysis. Estimates were positive among children of mothers who experienced low stress (externalizing: ψ=1.45, 95% CI: 0.59, 2.31, p=0.001; internalizing: ψ=1.39, 95% CI: 0.48, 2.29, p=0.003) and attenuated among children of mothers who experienced high stress (externalizing: ψ=0.73, 95% CI: −0.13, 1.57, p=0.10; internalizing: ψ=0.61, 95% CI: −0.22, 1.39, p=0.14). Modeled continuously, the interaction was - 0.12 per one-unit increase in the latent stress score for externalizing problems (95% CI: −0.31, 0.07; interaction p=0.20) and −0.17 for internalizing problems (95% CI: −0.36, 0.01; interaction p 0.07). However, no interaction was statistically significant under any of the three parameterizations. Taken together, we found no consistent evidence that prenatal maternal psychosocial stress modified the association between the prenatal PAH mixture and child behavioral problems (Table S7-S8).

### Effect modification by child sex

We assessed whether the associations between the prenatal mixtures (PAH metabolite mixture, psychosocial stress mixture, and joint mixture) and CBCL scores differed by child sex (Figure 5; Table S9). During the preschool waves, all three mixtures were associated behavioral problems among boys but not among girls. For the joint mixture, a one quintile increase was associated with a 3.42 point (95% CI: 2.09, 4.69) increase in externalizing problems among boys compared with 0.74 point increase (95% CI: −0.67, 2.14) among girls (interaction p=0.004), and a 3.25 point (95% CI: 1.90, 4.51) increase in internalizing problems among boys compared with 0.67 (95% CI: −0.86, 2.19) among girls (interaction p=0.009). Among boys, emotional IPV was the single largest contributor to the preschool joint mixture effect for both outcomes, followed by 2-NAP and 1-NAP for internalizing problems and by psychological distress and 1-NAP for externalizing problems. Psychosocial factors accounted for the majority of the positive component weights among boys for both outcomes (Table S10).

**Figure 5.**
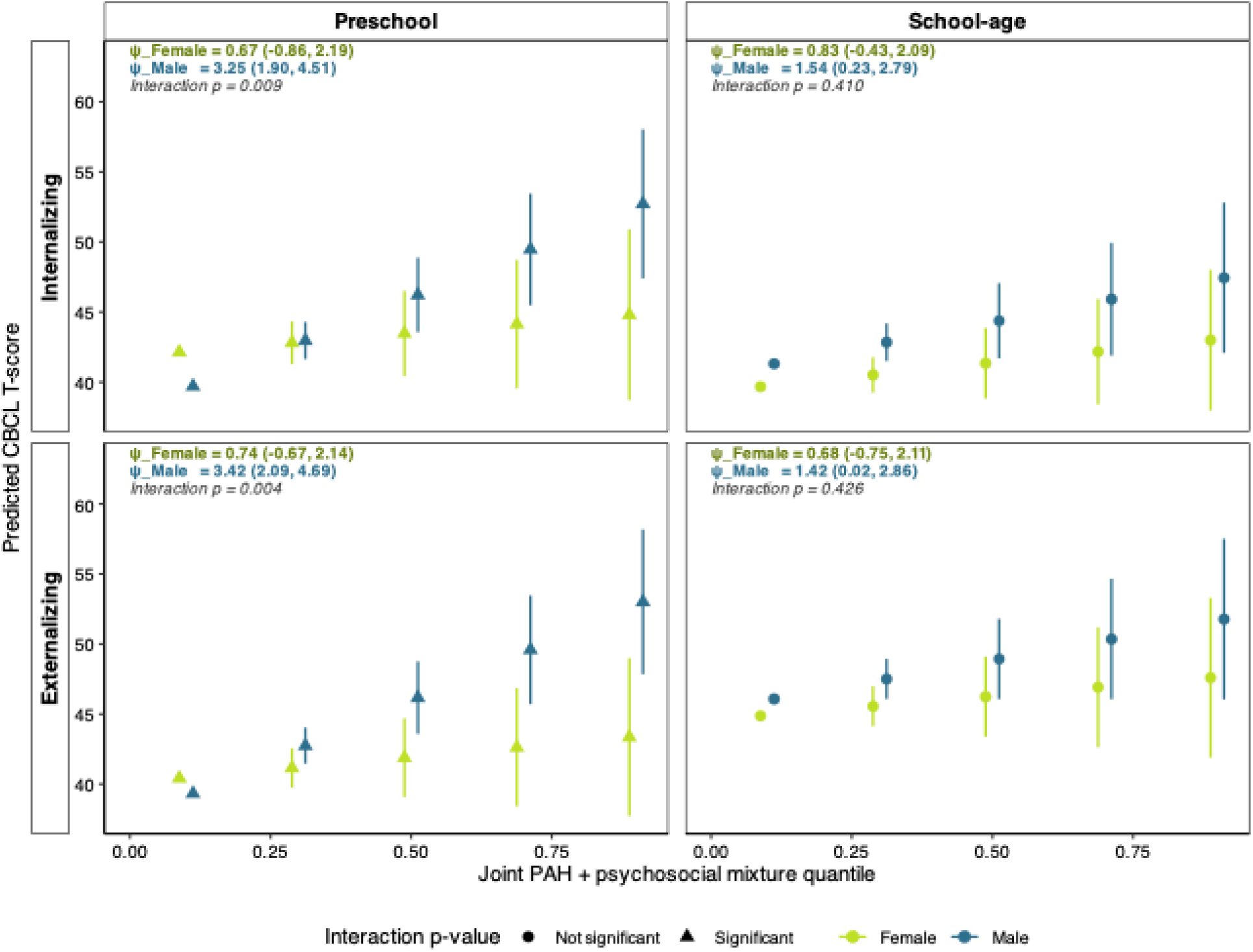
Effect measure modification by child sex of the association between the prenatal joint PAH and psychosocial mixture and child internalizing (top row) and externalizing (bottom row) problems at preschool (left column) and school-age (right column) windows. Stratum-specific QG-COMP estimates, stratified by child sex, female (green) and male (blue). Points give the predicted CBCL T-score at each quintile of the joint mixture and vertical lines are the corresponding 95% confidence interval. Point shape indicates whether the mixture by stress interaction p-value was statistically significant, with a triangle shape indicating statistical significance (p < 0.05). The estimated cumulative mixture effect (ψ) for each stratum, 95% confidence intervals, and interaction p-value are denoted within each panel. Ψ is the change in CBCL T-score per one quintile increase in all exposures simultaneously. All models were adjusted for self-reported race, child age at CBCL assessment, maternal age at enrollment, maternal smoking, maternal urinary creatinine concentration, and SES.

The same pattern of effect modification was observed for the psychosocial stress mixture and PAH metabolite mixtures during the preschool wave (Table S9-S10). For the psychosocial mixture, a one quintile increase in the mixture was associated with a 2.44 point (95% CI: 1.29, 3.54) increase in externalizing problems in boys compared with 0.29 (−0.95, 1.53) in girls (interaction p=0.013), and a 2.16 point (1.05, 3.22) increase in internalizing problems in boys compared with 0.40 (−0.88, 1.68) in girls (interaction p=0.045) during the preschool wave.

For the PAH metabolite mixture, a one quintile increase in the mixture was associated with a 1.43 point (95% CI: 0.55, 2.22) increase in internalizing problems in boys compared with 0.39 (−0.49, 1.28) in girls (interaction p=0.034). There was no statistically significant interaction in association with externalizing problems. During the school-age waves, no interaction with child sex was observed for any mixture (Table S9-S10).

### Quantitative bias analysis for exposure measurement error

Published ICCs for repeated urinary PAH metabolites across pregnancy were moderate and consistent between the two largest cohorts, falling within 0.04 of one another for every metabolite measured (Table S11). Correcting the single-metabolites estimates for measurement error scaled the effect estimates by a factor of 1.7 to 2.5 across the 0.40 to 0.57 ICC range reported in those cohorts (Table S12; Figure S10). Across the full range of ICCs, the bias-adjusted estimates exceeded the observed estimates in magnitude, indicated that unless the true ICC is 1, the reported single-metabolite associations are attenuated regardless of the reliability value assumed. Assuming an ICC of 0.50, for example, 2-FLUO was associated with a 2.98 point increase in externalizing problems (95% CI: 0.74, 5.20; vs. observed estimate of 1.49 [0.37, 2.60]), 1-NAP with a 2.28 point increase (0.42, 4.14 vs. 1.14 [0.21, 2.07]), and 1-PYR with a 2.10 point increase (0.28, 3.90 vs. 0.96 [0.07, 1.84]) per IQR increase in the natural log-transformed metabolite.

## Discussion

In this South African birth cohort, we found that the joint exposure to prenatal urinary PAH metabolites levels and psychosocial stressors was associated with child behavioral problems in the first decade of life, and that the two domains followed different developmental patterns. Associations with prenatal PAH metabolite levels, both individually and as a mixture, were confined to the preschool waves and were null by school age. In contrast, associations with psychosocial factors were of similar magnitude across both developmental windows. The joint effect of PAH metabolite levels and psychosocial factors produced larger associations than either domain alone, with psychosocial factors contributing most to the total effect. These associations differed by child sex, with all three mixtures associated with behavioral problems among boys, but not among girls during the preschool waves. We found no consistent evidence that these associations differed by prenatal maternal psychosocial stress. The remaining chemical classes, including pesticides, bisphenols, parabens, UV filters, and other biocides, were not associated with increased behavioral problems. Taken together, these findings suggest that prenatal PAH exposure and psychosocial stress are associated with child behavior through partly distinct developmental courses, and that the burden falls disproportionately on boys in early childhood.

We identified a contrast between the two exposure domains across development. Levels of three PAH metabolites, 2-FLUO, 1-NAP, and 1-PYR, were individually associated with externalizing problems during the preschool waves, and the PAH metabolite mixture was associated with both outcomes in this window. Neither the individual metabolites nor the mixture was associated with either behavioral outcome during the school-age waves. Psychosocial factors behaved differently. The latent stress score and the psychosocial mixture were associated with both outcomes in both developmental windows, and at a comparable magnitude. Prenatal chemical exposure may act on a narrower window of susceptibility, since the developing brain is most vulnerable to neurotoxic exposures during the periods of rapid growth in gestation and early life^63,64^, with its behavioral signature diluted as the postnatal environment accumulates. Psychosocial adversity measured in pregnancy, by contrast, may reflect a family environment that persists across childhood and continues to shape behavior. For example, in a meta-analysis of 76,514 children from seven European cohorts, associations between prenatal maternal depression and child behavioral problems were largely independent of pre-pregnancy depression but were partly mediated by continuing postnatal maternal depression^65^. Modeling time-varying postnatal exposures directly is an important direction for future work in this cohort.

Previous studies with repeated assessments of child behavior have reported persistence of the PAH metabolite level association over time, rather than attenuation. For example, in the Columbia Center for Children’s Environmental Health (CCCEH) cohort, children whose mothers had detectable PAH-DNA adducts levels showed almost no improvement in emotional self-regulation in children aged 3 and 11 years, while those who did not have detectable adducts levels improved substantially^66^. In the same cohort, higher prenatal PAH exposure, assessed by personal air monitoring and PAH-DNA adducts measurements, was associated with anxious/depressed and attention problems on the CBCL at 6 to 7 years of age^67^. Prenatal PAH-DNA adduct levels were associated with ADHD behavior problems at 9^68^ and in analyses combining PAH adduct levels with social adversity, associations were reported with attention and thought problems at age 11^69^. Our findings in the DCHS are not directly comparable to the CCCEH study. CCCEH was conducted in New York City, a high-income urban setting, whereas DCHS is set in a peri-urban region of Cape Town in a low- and middle-income country. Therefore, these cohorts differ in exposure sources, prevalence and intensity of exposure, socioeconomic context, and the broader postnatal environment children subsequently experience. Additionally, these studies characterized exposure using personal air monitoring and PAH-DNA adduct measurements rather than urinary PAH metabolite levels, which may capture different pathways and routes^70,71^.

No prior studies have assessed the association between prenatal urinary PAH metabolite levels and child psychopathology or behavioral outcomes across 10 years of childhood. A few prior studies conducted in the mainland US, Puerto Rico, and China, have assessed associations with neurodevelopmental outcomes, but their findings were mixed ^72–74^. For example, in the PROTECT cohort, they found lower odds of neurodevelopmental delay with higher prenatal urinary PAH metabolite levels from 12 to 48 months^74^. In the Tianjin birth cohort study, they did not identify any statistically significant associations with the Griffiths Development Scale at 6 to 12 months^73^.

Consistent with previous work in the DCHS, psychosocial factors contributed more to the joint mixture effect than environmental exposures^28,29,75,76^. When PAH metabolite levels and psychosocial factors were modeled together, the joint mixture was associated with both outcomes during the preschool waves, and the magnitude of this association was larger than that of either the PAH metabolite-only or the psychosocial stress-only mixture. A test of interaction between PAH metabolite levels and psychosocial stress provided no evidence that psychosocial factors modify the associations between PAH metabolite levels and child behavior. This suggests that PAHs and psychosocial factors contribute additively rather than synergistically to child behavior. Therefore, our findings point to a cumulative burden of two independent exposure domains where children exposed to both psychosocial stress and PAHs experience additive increases in child psychopathology.

The dominance of psychosocial factors may reflect the particularly high prevalence and severity of psychosocial burden in this South African population, where rates of depression, traumatic life events, and IPV are elevated in comparison to higher-income countries^42,47,77,78^. Two previous studies in this cohort identified a similar pattern using different exposure measures. Prenatal indoor air pollution and psychosocial factors were jointly associated with trajectories of internalizing and externalizing problems from 24 to 60 months^28^ and pregnancy was identified as the sensitive period for indoor air pollution, with evidence of a synergistic effect between air pollution and psychosocial factors in the prenatal window^29^. Taken together, these findings reinforce the preschool development window as a particularly sensitive window during which child behavioral is susceptible to environmental exposures and that psychosocial stress in combination with environmental exposures play a larger role in child behavioral development than either exposure alone.

The absence of associations for the remaining chemical classes is unlikely to reflect insufficient exposure or poor detection in our cohort. For example, prenatal pesticide metabolite concentrations in the DCHS exceeded those among pregnant women in the Center for the Health Assessment of Mothers and Children of Salinas (CHAMACOS) Study cohort of Mexican-American farmworkers in California’s Salinas Valley and in the National Health and Nutrition Examination Survey (NHANES 2003-2004) of the general US population [add refs]. Organophosphate metabolite concentrations in DCHS exceeded both: DMP was detected in 98% of participants, compared with 72% in CHAMACOS and 44% in NHANES, with a DCHS geometric mean (10.85 ng/mL) approximately six-fold higher than CHAMACOS (1.71 ng/mL). The high prenatal pesticide metabolite concentrations in this cohort likely reflect widespread agricultural pesticide use in the Western Cape, which is a major fruit- and wine-producing area where pesticide drift from vineyards into neighboring residential areas has been documented ^79^. Nevertheless, our findings add to the sparse literature on prenatal environmental exposures and child behavioral outcomes in LMIC settings.

DCHS PAH metabolite levels were broadly comparable to non-smoking US populations for naphthalene, fluorene, and phenanthrene metabolites, but 1-PYR levels were two to five times higher (DCHS geometric mean 0.30 ng/mL vs. 0.06 to 0.13 ng/mL) than across seven sites in three US pregnancy cohorts^80^ and higher than among non-smoking NHANES participants (0.30 ng/mL vs 0.10 ng/mL)^81^. Within sub-Saharan Africa, however, our concentrations were not unusually high. In the only comparable African pregnancy cohort, in Nairobi, Kenya, where 97% of participants had urinary cotinine below 10 ng/mL, geometric mean concentrations exceeded ours for both 2-NAP (17.5 ng/mL vs 4.84 ng/mL) and 1-PYR (0.52 vs 0.30 ng/mL)^82^. These comparisons are approximate, as studies differ in how urinary dilution was handled, how non-smoking was defined, and in the number of metabolite isomers included for each parent compound.

Several biological mechanisms could explain PAH-specific associations in early childhood. PAHs cross the placenta^83^ and their metabolites have been shown to cross the fetal blood-brain barrier and reach the developing central nervous system^84^. They may exert their adverse effects through oxidative stress, dysregulation of glutamatergic and catecholaminergic neurotransmission, and epigenetic modifications^84,85^. Prior studies have also identified associations between prenatal air pollution and brain-derived neurotrophic factor, dopaminergic signaling, and hippocampal structure^64,86^.

In this setting, the dominant PAH sources are likely to be combustion related, including biomass and paraffin fuel used for cooking and heating and vehicle exhaust. PAHs are produced when organic materials such as fossil fuels, wood or tobacco are burned. In indoor air, emissions from cooking, tobacco smoking, and moth repellents are expected to be the predominant sources^87^. Paraffin remains the primary cooking fuel for approximately two million South Africans, burnt in poorly made stoves that contribute to household air pollution^88^. In this cohort, indoor benzene concentrations measured during pregnancy exceeded ambient standards and were higher in homes using fossil fuels for cooking^89^. Previous studies have found that 1-NAP and 2-FLUO levels rise sharply with exposure to cooking and grilling smoke^90^, 1-PYR levels have been associated with dietary sources in addition to airborne sources, while 2-NAP has been associated with tobacco smoke^71,91^. As prenatal tobacco exposure was common in this cohort, we adjusted all models with PAH metabolites for cotinine-defined smoking categories to identify associations that are not explained by tobacco smoke as a source. In a prior study in this cohort, indoor levels of benzene and toluene were associated with increasing internalizing trajectories across pre-school^28^. Convergence of these findings across two exposure assessment methods—indoor air sampling and urinary metabolite biomarkers—strengthens the evidence that prenatal combustion-related exposure is associated with child behavior problems, particularly during preschool age.

Associations with all three mixtures (PAH-only, psychosocial only, or joint) differed by child sex during the preschool waves, with each mixture associated with behavioral problems among boys but not among girls. No interaction with child sex was observed during the school-age wave. These findings are consistent with previous studies that the male fetus may be more vulnerable to prenatal insults^20,21^. Sex-specific associations with prenatal PAH exposure have been largely inconsistent. In the Tianjin birth cohort, the association between urinary PAH metabolite mixture levels and social and language development were present among girls and not boys^73^. However, this study assessed neurodevelopmental domains at 6 to 12 months of age, earlier than our timepoints. Sex-specific associations have been reported more consistently for prenatal psychosocial stress than for prenatal chemical exposures. Prior studies have reported higher externalizing behavior among boys exposed to prenatal stress, while reviews have proposed that prenatal psychosocial stress contributes to greater female vulnerability to internalizing problems^92–94^. Our findings are consistent with this pattern for externalizing problems, but not for internalizing problems, as boys in our cohort were more affected for both outcomes, which suggests general male susceptibility during the preschool years.

The chemicals evaluated here have short biological half-lives, so a single urine sample collected during pregnancy likely characterizes recent rather than average gestational exposure, introducing classical, non-differential measurement error that biases effect estimates toward the null ^95,96^. In a multi-dimensional bias analysis sweeping across the range of ICCs previously reported for urinary PAH metabolites during pregnancy, we found that the observed effect estimates were consistently underestimated across the range of tested ICCs. Because the ICC is a variance ratio that depends on the distribution of true exposure within a population, the values applied here, which were estimated in pregnancy cohorts in Puerto Rico^57^ and New York City^58^, may not transport to a South African cohort with potentially varying exposure sources. Although sweeping across the full published range of ICC mitigates this, the range remains an assumption rather than an internally derived estimate. Nevertheless, adjustment using external reliability estimates generally reduces bias relative to the uncorrected estimate even where transportability does not fully hold^97^. Reliability also differed by metabolite, which suggests that metabolite-specific associations in any single-sample study partly reflect differing measurement reliability rather than differing toxicity, an implication that has not previously been quantified in studies of prenatal PAH exposure and child behavior. Taken together, these findings indicate that the associations reported here are more likely underestimates than overestimates of the association with average gestational exposure, and that future work should report metabolite-specific reliability alongside effect estimates.

Despite its strengths, our study has several limitations. First, many of the metabolites evaluated can also be preformed in the environment, such that co-exposure may occur to both the active compound and its inactive metabolite. Additionally, variable urine dilution in a spot sample may not be adequately adjusted with creatinine. Further, three analytes had low detection frequencies (< 50%), and although concentrations below the LOD were imputed from a left-censored log-normal distribution and the less detected analytes were dichotomized in single-exposure models, low detection may reduce power and bias associations toward the null. Next, the CBCL instrument changes between the 60 and 78 month assessments, and although both versions yield standardized T-scores on a common scale, the composites comprise different syndrome scales, with the Attention Problems subscale contributing to externalizing problems on the CBCL/1.5-5 but not on the CBCL/6-18. We adjusted for CBCL version when modeling all six timepoints together and modeled the two windows separately, but the outcomes are not identical constructs across that boundary. Comparison with the prior PAH literature is also constrained by how exposure was assessed. Most previous studies of prenatal PAH exposure and child behavior estimated exposure from personal air monitoring, which indexes the inhaled fraction, or from PAH-DNA adducts, which may reflect the biologically effective dose reaching the fetus. Urinary metabolites instead reflect internal dose from inhalation, diet, and dermal contact combined. In one study of pregnant women, urinary metabolite levels correlated with their airborne parent compounds only for only for phenanthrene and fluorene^71^. The three approaches likely do not capture the same exposure pathways, and thus estimates are not directly comparable. Differences between our findings and prior literature may therefore reflect the differences in exposure metric, outcome instrument, age at assessment, and the larger socioeconomic and geographic context of the population. Finally, because our exposures and central hypothesis concerns prenatal programming and we did not model concurrent postnatal exposures, we cannot determine whether the attenuation of prenatal exposure associations in school-age reflects an improving concurrent environment or the potential waning of prenatal effects over development. Modeling time-varying postnatal exposure directly is an important direction for future work in this cohort.

This study leverages rich data from a well-characterized, prospective birth cohort from a peri-urban South African community, a population that is both underrepresented in epidemiologic research and disproportionately burdened by environmental and psychosocial exposures. To our knowledge, this is the first study to examine prenatal urinary PAH metabolite levels in relation to child behavior in a sub-Saharan African cohort, and the first to do so with repeated outcome assessments spanning both the preschool and school-age years. The repeated assessment of child behavior at six timepoints through 10 years of age allowed us to assess whether chemical and psychosocial exposures follow different developmental courses. Additionally, the broad, multi-class chemical panel further allowed us to identify which exposure classes are associated with psychopathology. Future studies should replicate these findings in other LMIC cohorts, extend follow-up through adolescence, and work toward capturing the full exposome to more comprehensively characterize cumulative exposure burden in overburdened communities. Together, these findings advance our understanding of how environmental toxicants and psychosocial stressors shape child behavioral development across childhood and may ultimately contribute to environmental justice initiatives aimed at reducing health disparities in vulnerable and at-risk populations.

## Data Availability

The Drakenstein Child Health Study is committed to the principle of data sharing. De-identified data will be made available to requesting researchers as appropriate. Requests for collaborations are welcome. More information can be found on our website [http://www.paediatrics.uct.ac.za/scah/dchs].

## Funding and Acknowledgements

The authors would like to thank the study and clinical staff at Paarl Hospital, Mbekweni and TC Newman clinics, as well as the CEO of Paarl Hospital, and the Western Cape Health Department for their support of the study. The authors also thank the families and children who participated in this study.

The Drakenstein Child Health Study was funded by the Gates Foundation (OPP 1017641 and OPP1017579), the Wellcome Trust (221372/Z/20/Z), with additional support for this work from the South African Medical Research Council and the National Research Foundation of South Africa. The National Institutes of Health/National Institute of Environmental Health Sciences has awarded a grant to the Mount Sinai HHEAR Targeted Analysis Laboratory (U2CES026561, PI: Robert O. Wright) for the laboratory analysis of urine creatinine and urinary chemical metabolites.

Research reported in this publication was supported by the National Institute of Environmental Health Sciences of the National Institutes of Health (NIH) under award numbers T32ES012870 (S.A.) and F31ES037540 (S.A.). Approximately 65% of total project costs were financed with federal funds in the amount of $150,000. The remaining 35% of the total project costs were financed by non-governmental sources in the amount of approximately $80,000. The NIH had no further role in study design; in the collection, analysis and interpretation of data; in the writing of the report; and in the decision to submit the paper for publication. The content is solely the responsibility of the authors and does not necessarily represent the official views of the NIH. This manuscript is subject to the NIH Public Access Policy. Through acceptance of this federal funding, NIH has been given a right to make this manuscript publicly available in PubMed Central upon the Official Date of Publication, as defined by NIH.

The authors also wish to disclose that a preprint version of this manuscript is available on medRxiv.

