## Supplemental Figures for "Environmental Chemical and Psychosocial Stress Exposures during Pregnancy and Child Behavioral Problems through 10 Years of Age in a South African Birth Cohort"

### Supplementary Figures

Sarina Abrishamcar^1^, Stephanie M. Eick^1,2^, Todd M. Everson^1,2^, M. Daniele Fallin^1^, Shakira F. Suglia^1^, Jasmin Chovatiya^3^, Ravikumar Jagani^3^, Robert O. Wright^3^, Syam S. Andra^3^, Dana Boyd Barr^2^, Nadia Hoffman^4,5^, Heather J. Zar^6^, Dan J. Stein^†4,5^, Anke Hüls^1,2,7^ *

^1^Department of Epidemiology, Rollins School of Public Health, Emory University, Atlanta, Georgia, 30322, USA

^2^Gangarosa Department of Environmental Health, Rollins School of Public Health, Emory University, Atlanta, Georgia, 30322, USA

^3^Department of Environmental Medicine, Icahn School of Medicine at Mount Sinai, New York, New York, 10029, USA.

^4^Dept of Psychiatry & Mental Health and Neuroscience Institute, University of Cape Town, Cape Town, 7700, South Africa

^5^South African Medical Research Council (SAMRC) Unit on Risk and Resilience in Mental Disorders, University of Cape Town, Cape Town, 7700, South Africa

^6^Department of Paediatrics and Child Health, SA-MRC unit on Child & Adolescent Health, Red Cross War Memorial Children’s Hospital, University of Cape Town, Cape Town, 7925, South Africa

^7^Department of Biostatistics and Bioinformatics, Rollins School of Public Health, Emory University, Atlanta, Georgia, 30322, USA

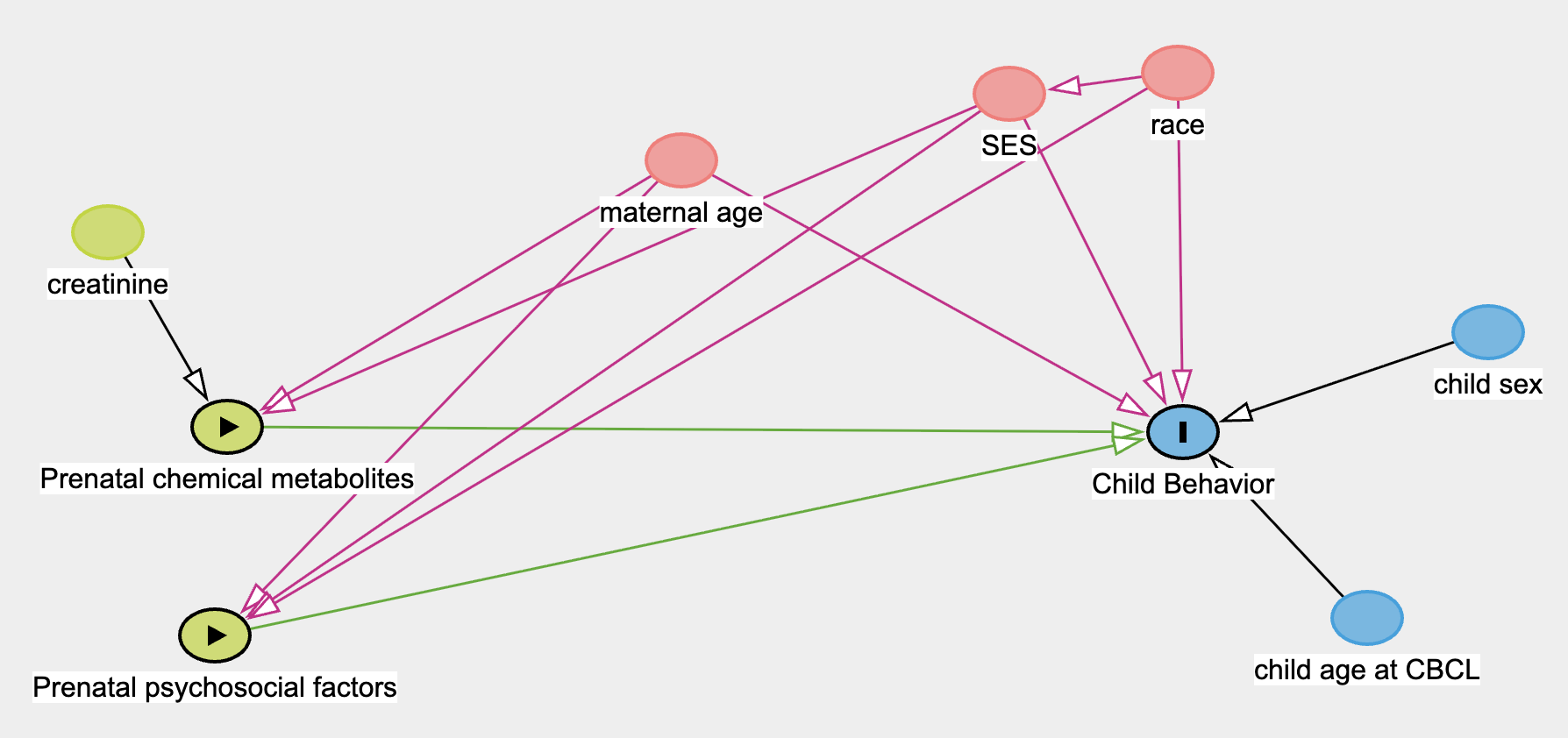
**Figure S1:** Directed acyclic graph for confounding and covariate adjustment.

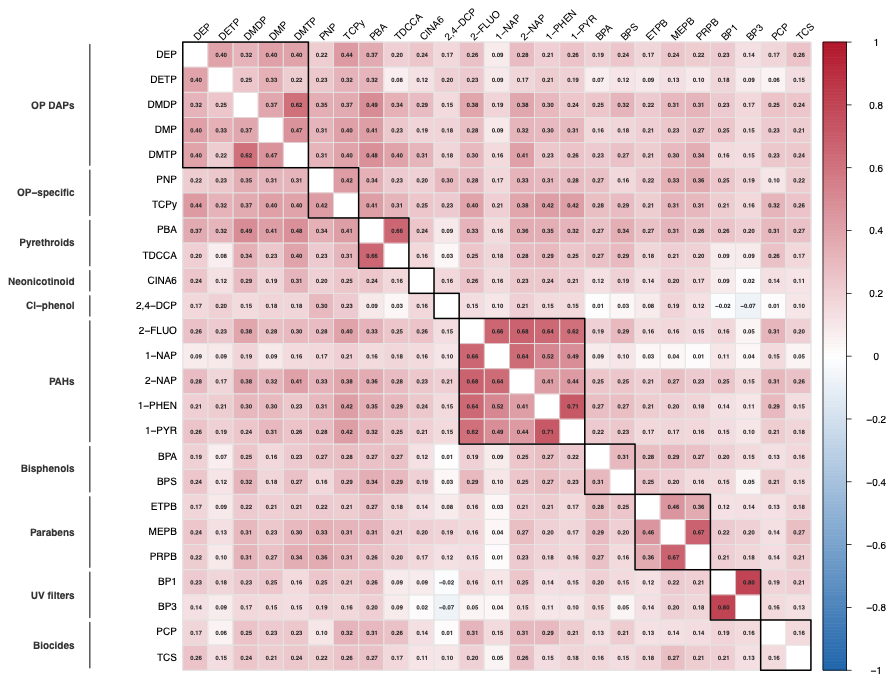
**Figure S2:** Pairwise Pearson correlations between prenatal urinary pesticide metabolite concentrations. Correlation coefficients (r) are displayed within each cell, with color intensity reflecting the strength and direction of the correlation. Chemical classes are boxed.
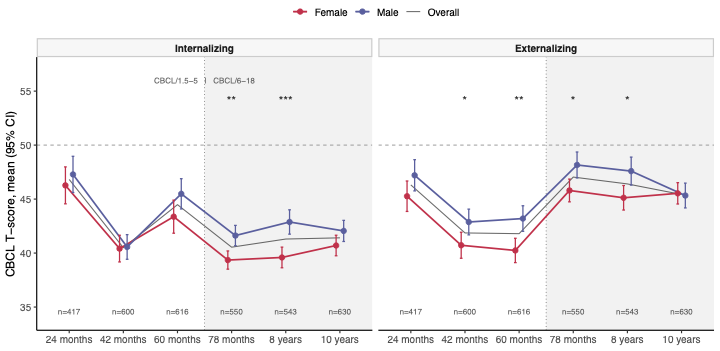
**Figure S3: Observed CBCL internalizing (left) and externalizing (right) T-scores across the six assessment waves, by child sex**. Points and vertical lines give the mean and 95% confidence interval among children with a non-missing score at that wave, shown separately for female (pink) and male (purple) children. The thin gray line is the overall mean. Sample sizes at each wave are given along the bottom axis. Estimates are from observed data, before multiple imputation. The horizontal dashed line at T = 50 marks the normative mean of the CBCL standardization sample. Because T-scores are already standardized for age and sex, a difference between the two series indicates that boys in this cohort scored higher relative to US norms than girls did. The vertical dotted line marks the change in instrument between the 60 and 78 month assessments, from the CBCL/1.5-5 to the CBCL/6-18, which differ in both items and norming sample. Asterisks denote the between-sex difference at each wave from Welch t-tests, adjusted across the 12 waves by scale comparisons using the Benjamini-Hochberg procedure: * q < 0.05, ** q < 0.01, *** q < 0.001.
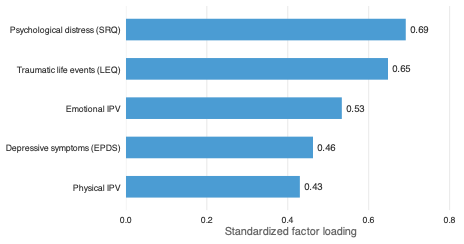
**Figure S4.** Standardized factor loadings from the confirmatory factor analysis (CFA) of the latent prenatal psychosocial stress construct. Standardized factor loadings reflect the strength of each indicator's contribution to the latent construct.

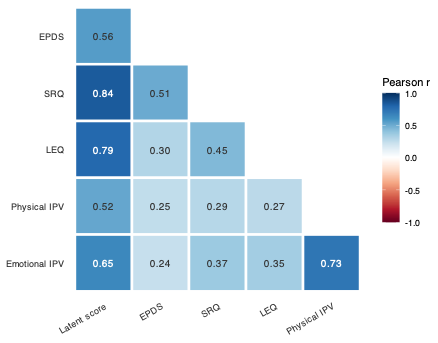

**Figure S5**. Pairwise Pearson correlations between prenatal psychosocial factor indicators and the latent psychosocial stress score. Correlation coefficients (r) are displayed within each cell, with color intensity reflecting the strength and direction of the correlation.

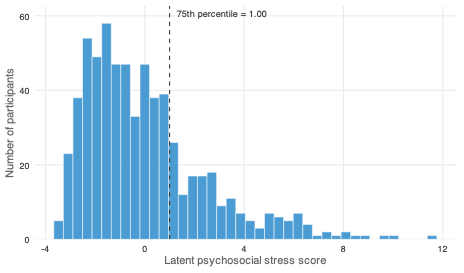

**Figure S6:** Distribution of the latent prenatal psychosocial stress score estimated from the confirmatory factor analysis model. The dashed vertical line indicates the 75th percentile (score = 1.00), which was used as the threshold to define high versus low psychosocial stress groups in effect measure modification analyses.
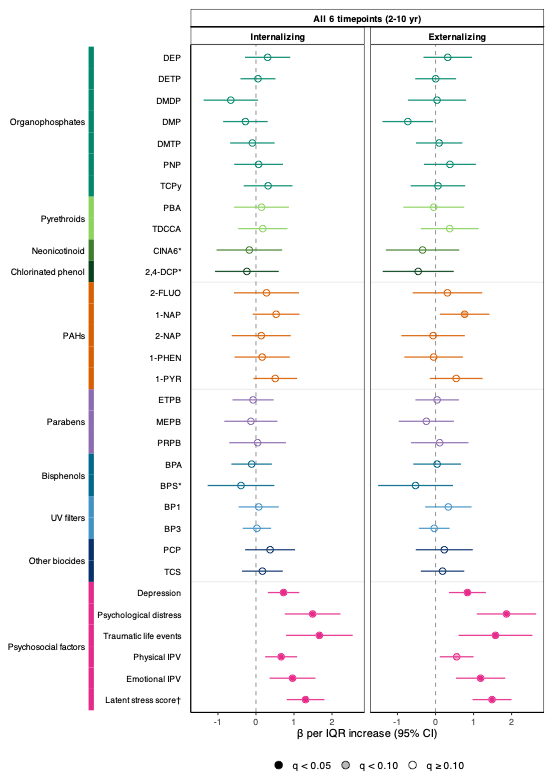
**Figure S7. Single-exposure associations between prenatal environmental or psychosocial exposures and child internalizing and externalizing problems across all six timepoints from 2 to 10 years.** Estimates are from GEE models with an exchangeable working correlation, fit across 35 multiply imputed datasets and pooled by Rubin’s rules. Point estimates denote the change in CBCL T-score per one interquartile range (IQR) increase in the exposure, with horizontal lines denoting the corresponding 95% confidence intervals. The colored bar groups the exposure into ten exposure classes. The BH-FDR correction was applied across all exposures within each outcome and developmental window. Point estimate fill indicates the FDR-corrected q-value: filled q <0.05, gray q >0.10, and open q ≥ 0.10.

All models were adjusted for self-reported race, child sex, child age at CBCL assessment, CBCL version, maternal age at enrollment, and SES. Models for urinary chemical metabolites were additionally adjusted for maternal urinary creatinine concentration and models for PAH metabolites were further adjusted for maternal smoking.

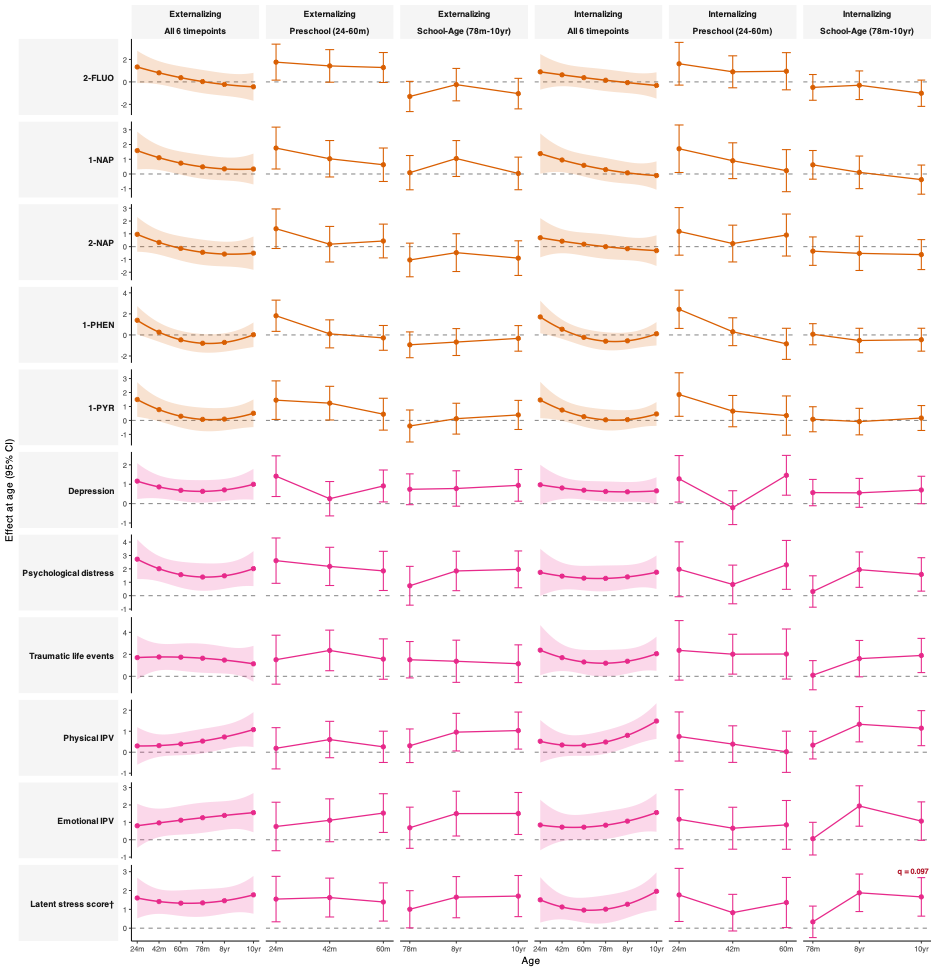
**Figure S8:** Time-varying associations between prenatal PAH metabolites and psychosocial stress and child internalizing and externalizing problems from 2 to 10 years of age. Rows show the 11 exposures carried forward from the single-exposure models. Columns show externalizing problems and internalizing problems each across all six assessments, the preschool window, and the school-age window. Points and intervals denote the change in CBCL T-score per IQR increase in the exposure at that age, with 95 Cis, estimated from GEE models including exposure by age and exposure by age-squared interaction terms and fit across 35 multiply imputed datasets. Models with D2 interaction q-values < 0.10 are annotated.

All models were adjusted for self-reported race, child sex, child age at CBCL assessment, maternal age at enrollment, and SES. Models that included all six assessments were additionally adjusted for CBCL version. Models for PAH metabolites were additionally adjusted for maternal urinary creatinine concentration and maternal smoking.

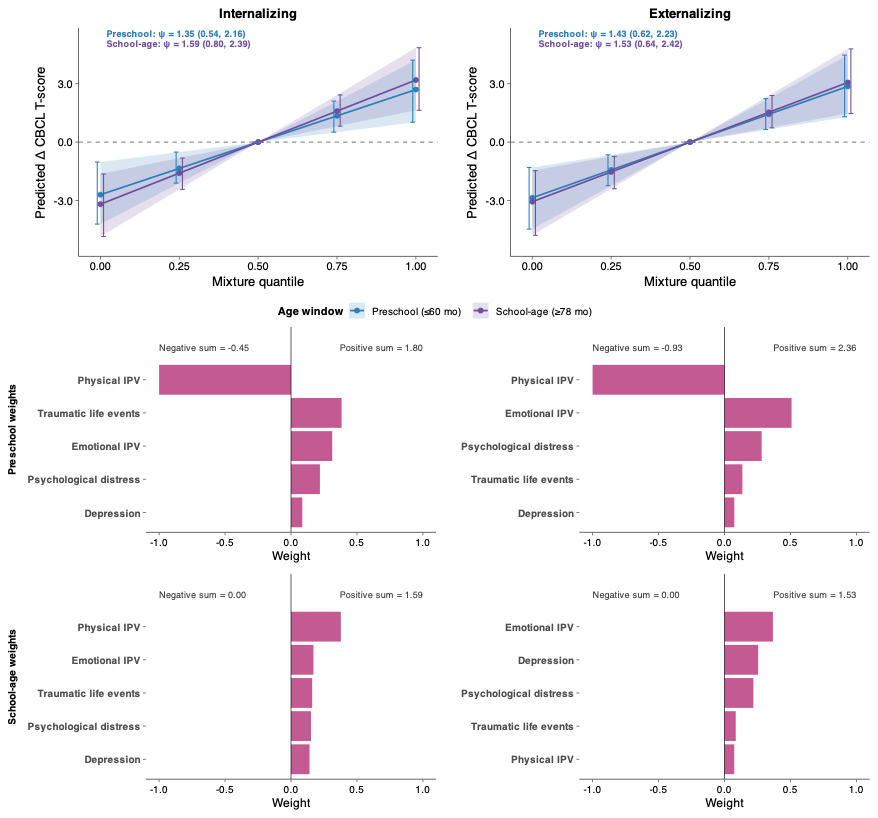
**Figure S9:** Quantile g-computation (QG-COMP) mixture associations between the prenatal psychosocial stress mixture and child internalizing and externalizing behavior at the preschool and school-age windows. Columns show internalizing (left) and externalizing (right) problems. **Top row:** QG-COMP marginal dose response curves for preschool (blue) and school-age (purple) developmental windows, showing the predicted change in CBCL T-score per one quintile increase in all exposures simultaneously, relative to the middle quintile, along with 95% confidence intervals. The estimated cumulative mixture effect (ψ) and 95% confidence intervals are denoted. **Middle and bottom row**: QG-COMP mixture weights for the preschool and school-age windows, indicating the relative contribution and direction of each exposure to the overall mixture effect. Weights are normalized and therefore sum to 1 within each direction. The positive sum and negative sum are the summed effects of the positively and negatively weighted components, with their net equivalent to ψ.

All models were adjusted for self-reported race, child sex, child age at CBCL assessment, maternal age at enrollment, and SES.

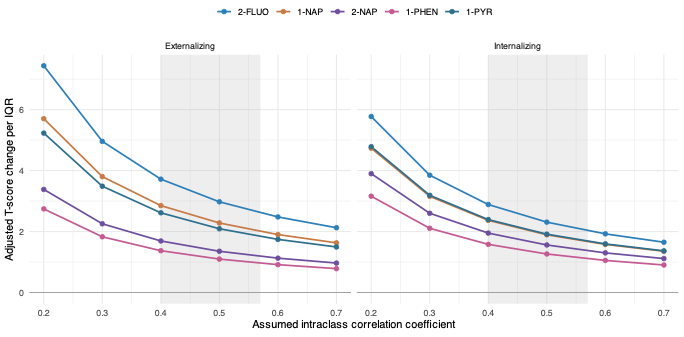

**Figure S10:** Bias-adjusted associations between prenatal urinary PAH metabolite levels and child behavioral problems across a grid of assumed intraclass correlation coefficients (ICCs). **Left:** Externalizing problems during the preschool waves. **Right:** Internalizing problems during the preschool waves. Points denote the bias-adjusted change in CBCL T-score per IQR increase in each metabolites, calculated by dividing the observed effect estimate by the assumed ICC. The shaed band spans 0.40 to 0.57, the range of per-metabolite values reported by the two largest pregnancy cohorts.

All models were adjusted for self-reported race, child sex, child age at CBCL assessment, matenral age at enrollment, SES, maternal smoking, and maternal urinary creatinine concentration.
